# Leveraging global multi-ancestry meta-analysis in the study of Idiopathic Pulmonary Fibrosis genetics

**DOI:** 10.1101/2021.12.29.21268310

**Authors:** Juulia J. Partanen, Paavo Häppölä, Wei Zhou, Arto Aleksanteri Lehisto, Mari Ainola, Eva Sutinen, Richard J Allen, Amy D Stockwell, Justin M Oldham, Beatriz Guillen-Guio, Carlos Flores, Imre Noth, Brian L Yaspan, R. Gisli Jenkins, Louise V Wain, Samuli Ripatti, Matti Pirinen, Global Biobank Meta-analysis Initiative (GBMI), Riitta Kaarteenaho, Marjukka Myllärniemi, Mark J Daly, Jukka T. Koskela

## Abstract

The research of rare and devastating orphan diseases such as Idiopathic Pulmonary Fibrosis (IPF) has been limited by the rarity of the disease itself. The prognosis is poor – the prevalence of IPF is only ∼4-times the incidence of the condition, limiting the recruitment of patients to trials and studies of the underlying biology of the disease. However, global biobanking efforts can dramatically alter the future of IPF research.

Here we describe the largest meta-analysis of IPF, with 8,492 patients and 1,355,819 population controls from 13 biobanks around the globe. Finally, we combine the meta-analysis with the largest available meta-analysis of IPF so far, reaching 11,160 patients and 1,364,410 population controls in analysis.

We identify seven novel genome-wide significant loci, only one of which would have been identified if the analysis had been limited to European ancestry individuals. We observe notable pleiotropy across IPF susceptibility and severe COVID-19 infection, beyond what is known to date. We also note a significant unexplained sex-heterogeneity effect at the strongest IPF locus *MUC5B*.

## Introduction

Idiopathic pulmonary fibrosis (IPF) is a chronic, progressive fibrotic disease of the lungs. It has no known etiology and pathogenesis, a poor prognosis, and limited treatment options. The prevailing model of IPF pathogenesis suggests recurrent epithelial injury followed by aberrant repair and dysregulated interstitial matrix deposition with cell senescence playing an important role in promoting lung fibrosis^1^.

As IPF has, by definition, no identifiable cause, genome-wide approaches are especially attractive as they may provide insight into underlying causes, pathogenesis, and might potentially reveal novel therapeutic avenues. Genome-wide association studies (GWAS) of IPF have thus far reported at least 23 associated loci^2–11^ highlighting genes involved with telomere maintenance^12^, cell adhesion, airway clearance, and innate immunity. These studies have mainly been restricted to individuals of European descent and common variants, and have identified few associations to conclusively functional variants. The most recent and largest meta-analysis concluded that IPF is highly polygenic with a significant number of associated variants remaining to be identified^2^. In addition, considerable genetic overlap between IPF and severe coronavirus disease 2019 (COVID-19) has been reported^13–16^.

To further explore the genetics of IPF susceptibility, we performed the first multi-ancestry study on the genetics of IPF in six populations, altogether comprising a 4-fold increase in the number of patients compared to the largest IPF study to date, via meta-analysis of the Global Biobank Meta-Analysis Initiative (GBMI) with the most recent published IPF study^2^. This allowed assessment of heterogeneity of effects over different ancestries and IPF diagnosis ascertainment between biobank and clinical cohort studies. With the increase in power, we were able to study population specific, rare, and sex-dependent variant effects. A notable fraction of proposed loci have previously been associated with lung function measured by spirometry. Fine-mapping the identified loci in the Finnish population, making use of reduced allelic heterogeneity of a population isolate, identified a functional causal variant in the previously reported *KIF15* locus. We describe significant pleiotropy between IPF and coronavirus disease 2019 (COVID-19), beyond what is known to date. Finally, we reveal significant sex-based heterogeneity at *MUC5B*, the strongest genetic risk factor of IPF.

## Results

### Multi-ancestry meta-analysis reveals 7 novel IPF loci

The GBMI IPF meta-analysis consisted of 8,492 cases and 1,355,819 controls representing six ancestries (Table 1). While IPF prevalence and recruitment strategies varied greatly across contributing biobanks, the overall prevalence was 0.62% (Table S1). The GBMI IPF meta-analysis discovered 16 genome-wide significant loci, highlighting two potentially novel loci (Figure 1).

**Table 1.** Ancestries in the GBMI IPF meta-analysis. AFR = African/African American, AMR = Latino/Admixed American, EAS = East Asian, FIN = Finnish, NFE = Non-Finnnish European, SAS = South Asian. Biobanks: BioME, BioVU, Colorado Center for Personalized Medicine Biobank (CCPM), Michigan Genome Initiative (MGI), UCLA Precision Health Biobank (UCLA), and Mass General Brigham (MGB) in America, Biobank Japan (BBJ) and China Kadoorie Biobank (CKB) in East Asia, and Genes & Health (GNH), Estonian Biobank (ETB), FinnGen project, Trøndelag Health Study (HUNT), and UK Biobank (UKBB) in Europe.

| Ancestry | Biobanks | Number of IPF patients | Number of controls | Fraction of cases (%) |
| --- | --- | --- | --- | --- |
| NFE | BioVU, CCPM, ESTBB, HUNT, MGB, MGI, UCLA, UKBB | 5,229 | 750,630 | 0.692 |
| FIN | FinnGen | 1,514 | 306,063 | 0.492 |
| EAS | BBJ, CKB, UCLA | 1,210 | 254,409 | 0.473 |
| AMR | BioMe, UCLA | 319 | 14,452 | 2.160 |
| AFR | BioMe, UCLA | 169 | 8,368 | 1.980 |
| SAS | GNH | 51 | 21,897 | 0.232 |
| **6** | **13** | **8,492** | **1,355,819** | **0.622** |

**Figure 1.**
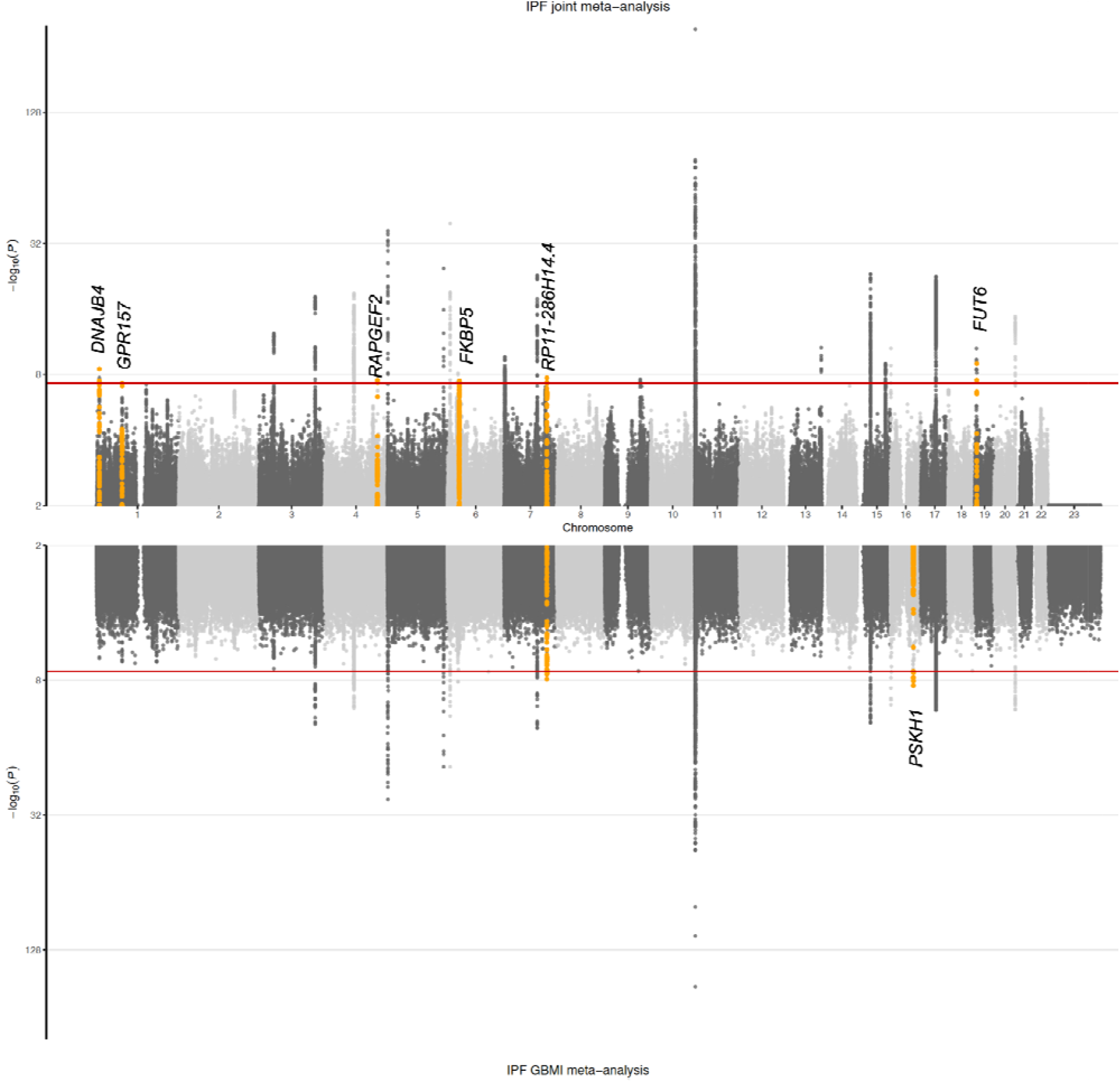
Genome-wide association results for IPF. Results from the joint meta-analysis are plotted in th top panel and results from the GBMI meta-analysis in the bottom panel. Novel associations are highlighted in orange and annotated with the closest gene (index variant and variants in LD r^2^ > 0.05 with it are highlighted, except for the *PSKH1* signal for which variants within a 1 Mb window are highlighted due to missing LD information).

We then meta-analyzed the GBMI data with the largest IPF meta-analysis ^2^ to date, later referred to as the Allen et al. study, increasing the number of cases and controls to 11,160 and 1,364,410, respectively. Altogether, the joint meta-analysis identified 25 independent IPF associated loci (Figure 1, Figure S1, Table S2). We report genome-wide significant results at 14/23 previously reported loci, with a further 7/23 showing consistent direction of effects at varying levels of significance (Table S3). The LD score regression intercept^17^ for the joint meta-analysis was not inflated (1.012) indicating independence of included studies (see Methods). Quantile-quantile plots for meta-analyses are available in Figure S2.

Beyond confirming nearly all of the previous signals, we identified seven potentially novel loci (Table 2, Figure 1). One locus was driven by rs539683219 at 16q22.1, an intronic *PSKH1* variant polymorphic only in the East Asian population. Highlighting the importance of cross-continental analysis, four out of the seven novel loci were mostly driven by non-European ancestry, when assessed by highest minor allele frequency at population level within the meta-analysis. Minor allele frequency enrichment within the meta-analysis compared to non-Finnish Europeans was over 1.5-fold for these four index variants. Moreover, if only the European populations (including non-Finnish Europeans and Finnish) were analyzed, only one of the seven loci reached genome-wide significance (Table S4).

**Table 2.**
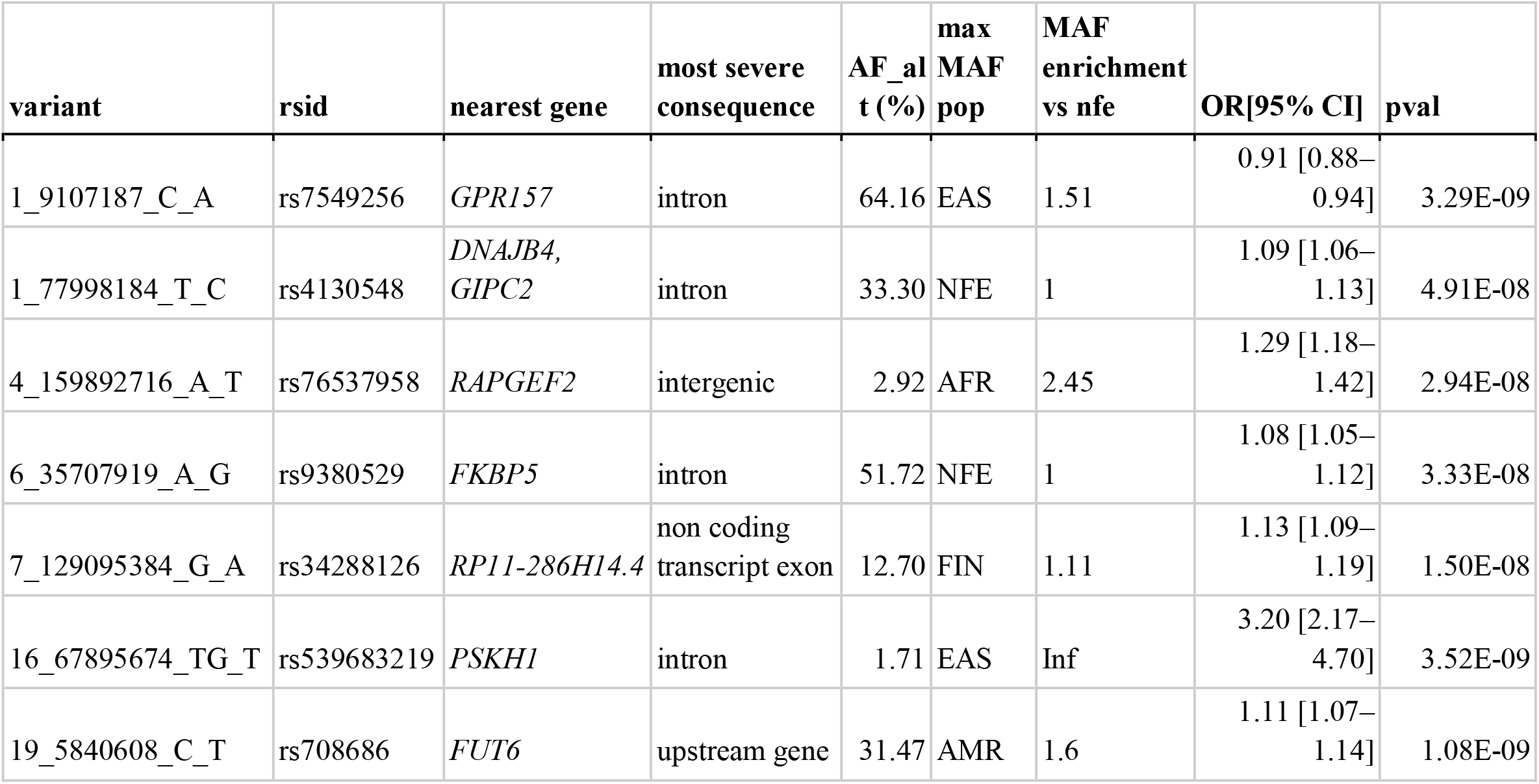
Novel associations from the GBMI and joint IPF meta-analyses. Nearest gene and most severe consequence information form Variant Effect Predictor (VEP), AF_alt = within the meta-analysis sample size weighted GRCh38 alternate allele frequency across studies included, max MAF population = population with highest minor allele frequency (MAF), MAF enrichment is calculated as highest MAF divided by MAF in the non-Finnish European population. AFR = African/African American, AMR = Latino/Admixed American, EAS = East Asian, FIN = Finnish, NFE = Non-Finnnish European, SAS = South Asian. Inf = Polymorphic in only one population.

| variant | rsid | nearest gene | most severe consequence | AF<sub>alt</sub> (%) | max MAF pop | MAF enrichment vs nfe | OR[95% CI] | pval |
| --- | --- | --- | --- | --- | --- | --- | --- | --- |
| 1_9107187_C_A | rs7549256 | *GPR157* | intron | 64.16 | EAS | 1.51 | 0.91 [0.88–0.94] | 3.29E-09 |
| 1_77998184_T_C | rs4130548 | *DNAJB4*,<br>*GIPC2* | intron | 33.30 | NFE | 1 | 1.09 [1.06–1.13] | 4.91E-08 |
| 4_159892716_A_T | rs76537958 | *RAPGEF2* | intergenic | 2.92 | AFR | 2.45 | 1.29 [1.18–1.42] | 2.94E-08 |
| 6_35707919_A_G | rs9380529 | *FKBP5* | intron | 51.72 | NFE | 1 | 1.08 [1.05–1.12] | 3.33E-08 |
| 7_129095384_G_A | rs34288126 | *RP11-286H14.4* | non coding transcript exon | 12.70 | FIN | 1.11 | 1.13 [1.09–1.19] | 1.50E-08 |
| 16_67895674_TG_T | rs539683219 | *PSKH1* | intron | 1.71 | EAS | Inf | 3.20 [2.17–4.70] | 3.52E-09 |
| 19_5840608_C_T | rs708686 | *FUT6* | upstream gene | 31.47 | AMR | 1.6 | 1.11 [1.07–1.14] | 1.08E-09 |

Further replication of the novel loci was attempted in two individual European ancestry cohorts (case count = 792 and 664), where six loci were polymorphic and imputed at high quality (minimum imputation R^2^ = 0.98). Three of the six potentially novel findings were replicated (at p-value < 0.01 and with consistent direction of effects, Table S5).

### Multiple variants associated with lung function and different organ manifestations

The Open Targets^18^ resource was used to assess previous findings for the proposed novel loci. Three of the seven novel loci have been previously implicated with lung function; at 6p21.31, the index variant rs9380529 in *FKBP5* was found in the 95% credible set for Forced Vital Capacity (FVC)^19^. The index variant for FVC (rs28435135, linkage disequilibrium, LD, r^2^ = 0.28) had a negative beta coefficient, i.e. decreasing effect on vital capacity, consistent with higher risk of IPF suggested in the present study. In addition to FVC, rs9380529 was in LD with the lead variant (and included in the 95% credible set) for trunk and leg fat percentages in UKB Neale v2 analysis (r^2^ = 0.64, http://www.nealelab.is/uk-biobank/).

*GPR157* at 1p36.22 has been associated with Forced Expiratory Volume in 1 second (FEV1) / FVC-ratio^20^, as the index variant rs7549256 from the current analysis was in LD (r^2^ = 0.55) with the reported index variant (no data on credible set or effect available). Rs7549256 was also in LD (r^2^ = 0.64) with the index variant for insulin-like growth factor 1 levels ^21^.

No previous associations were found for the intergenic rs76537958 at 4q32.1, while variants (LD with rs76537958: r^2^ < 0.1) in *RAPGEF2* have been associated with FVC in UKB Neale v2 analysis, childhood and lifetime pneumonia^22^. In addition, *RAPGEF2* has been reported as a shared risk factor for both IPF and chronic obstructive pulmonary disease (COPD) in a network analysis^23^.

At 1p31.1, rs4130548 has been implicated especially in Body Mass Index (BMI) and was the index variant of the association signal^24^. At 16q22.1, no previously reported associations were found for the EAS-specific intronic rs539683219 in *PSKH1*. At 19p13.3, the rs708686 upstream to *FUT6* has been previously associated with Carbohydrate Antigen 19.9 (CA-19.9)^25^, blood protein levels (FUT3)^26^, and gallstones ^26,27^, among others. At 7q32.1, the non-coding transcript exon variant rs34288126 has not been previously associated with any disease.

In contrast with other pulmonary diseases attributed to tobacco smoke exposure, such as COPD and lung cancer, no association signal was seen in the *CHRNA3/5* locus (OR[95% CI] = 1.05[1.02-1.08], p = 0.0034 for rs16969968), a known nicotine dependence locus^28^.

### Fine-mapping in the Finnish population suggests missense variant to be causal at *KIF15*

To identify potential causal alleles within the identified loci, we performed fine-mapping of all identified loci in FinnGen. This resulted in eight independent loci with suggested causal alleles (Table 3), while none of the novel loci were successfully fine-mapped (with good quality credible sets, i.e. minimum LD between variants r^2^ ≥0.25) in FinnGen.

**Table 3.** Fine-mapped good quality (minimum LD between variants r2 0.25) credible sets. PIP = posterior inclusion probability. Variants are reported based on the human genome reference sequence GRCh38.

| Locus | # variants<br>in credible<br>set | credible set<br>min LD ($r^2$) | highest PIP<br>variant | gene for<br>most severe<br>consequence | most severe<br>consequence | PIP |
| --- | --- | --- | --- | --- | --- | --- |
| 3q26.2 | 35 | 0.91 | 3:169759718:A:G | *ACTRT3* | downstream<br>gene | 0.080 |
| 3p21.31 | 22 | 0.48 | 3:44801967:G:T | *KIF15* | missense | 0.243 |
| 5p15.33 | 1 | 1.00 | 5:1272247:G:A | *TERT* | stop gained | 1.000 |
| 5p15.33 | 1 | 1.00 | 5:1279370:T:C | *TERT* | missense | 0.997 |
| 5q35.1 | 2 | 0.82 | 5:169588475:G:A | *SPDL1* | missense | 0.875 |
| 11p15.5 | 1 | 1.00 | 11:1219991:G:T | *MUC5B* | upstream gene | 1.000 |
| 11p15.5 | 2 | 0.34 | 11:1468491:G:A | *MOB2* | downstream<br>gene | 0.919 |
| 16p13.3 | 5 | 0.78 | 16:276685:G:A | *ARHGDIG* | intron | 0.618 |
| 17q21.31 | 2180 | 0.95 | 17:45753401_4575<br>3524del | *CRHR1* | intron | 0.003 |
| 20q13.33 | 17 | 0.51 | 20:63582806:G:A | *GMEB2* | downstream<br>gene | 0.077 |

Fine-mapping suggested deleterious coding causal variants at three loci. In addition to the previously reported coding variants in *TERT* and *SPDL1*^*4,6*^, fine-mapping identified a coding variant in *KIF15* (predicted missense, rs138043992, AF = 0.29%, OR[95% CI] = 1.71[1.39-2.10], PIP = 0.24), enriched 2.6-fold in the Finnish population compared to non-Finnish non-Estonian Europeans (NFEE, gnomAD v2.1.1) and predicted as probably damaging by Polyphen and deleterious by SIFT.

When a known locus near *KANSL1/MAPT* was assessed, the signal was fine-mapped to *CRHR1* (intronic rs1568002709, AF = 11.7%, OR[95% CI] = 0.62[0.53-0.73], PIP = 0.003). There was, however, little resolution in the locus due to its exceptional LD structure ^29^(Figure S1). Fine-mapping further suggested two independent signals at 11p15.5; the well-established *MUC5B* and an independent signal downstream to *MOB2*, see Supplementary Results for details.

### Pleiotropy of effects across COVID-19 severity

As considerable genetic overlap between IPF and severe COVID-19 caused by SARS-CoV-2 infection has been reported^13–16^, we assessed the shared genetic background of IPF and severe COVID-19 using the largest sample sizes available for both traits: the joint IPF meta-analysis reported here and the most recent COVID-19 Host Genetics Initiative (HGI) results (data release 6, previous release has been published^13^). We discovered that, in addition to the four previously reported loci (*MUC5B, DPP9, KANSL1/CRHR1*, and *ZKSCAN1*)^13,14,15,16^ associated with both IPF and COVID-19 hospitalization at a genome-wide level, three other genome-wide significant loci in the IPF meta-analysis passed the FDR-adjusted p-value threshold of 0.05 in the COVID-19 scan (7/25, 28%, Figure 2, Table 4). As previously reported, the effect of *MUC5B* was reversed: the strong, established risk allele in IPF is clearly protective for severe COVID-19 (OR = 0.89, p = 1.2E-8). The *ATP11A* locus also demonstrated opposite effects for the two traits, while for the rest of the loci the direction of effects was shared. While genome-wide associations for both IPF and COVID-19 hospitalization have been reported separately in the 17q21.31 locus, we noted a shared signal at the locus with very high LD between the index variants (r^2^ = 0.97). Secondly, six of the 17 loci from the COVID-19 hospitalization scan passed the FDR-adjusted p-value threshold of 0.05 in the IPF meta-analysis (35%, Table S6), suggesting further shared etiology at *CCHCR1, SLC22A31* and *TAC4*.

**Table 4.**
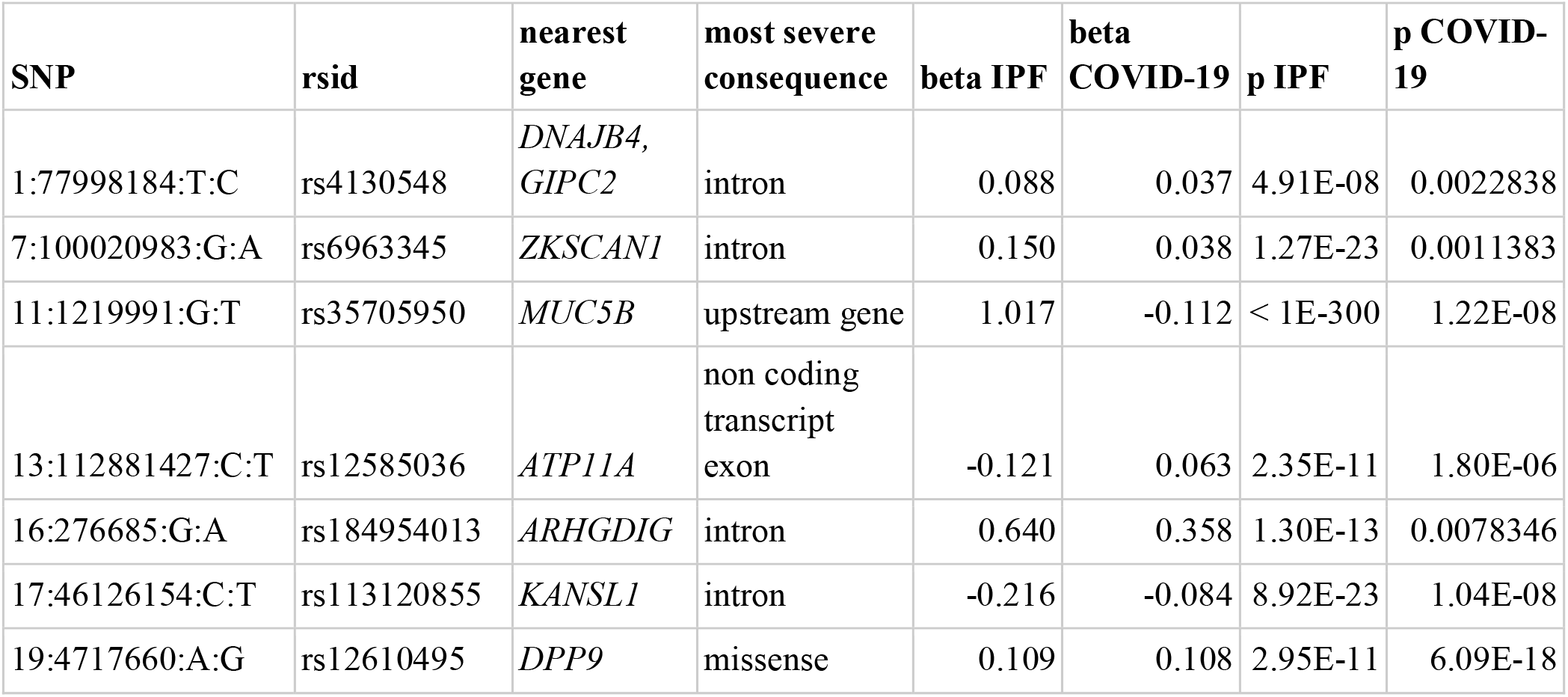
COVID-19 pleiotropy. Genome-wide significant index variants in joint IPF meta-analysis with FDR-adjusted p-value < 0.05 in COVID-19 hospitalization scan (n = 7 / 25). Variants are reported based on the human genome reference sequence GRCh38.

| SNP | rsid | nearest gene | most severe consequence | beta IPF | beta COVID-19 | p IPF | p COVID-19 |
| --- | --- | --- | --- | --- | --- | --- | --- |
| 1:77998184:T:C | rs4130548 | *DNAJB4*,<br>*GIPC2* | intron | 0.088 | 0.037 | 4.91E-08 | 0.0022838 |
| 7:100020983:G:A | rs6963345 | *ZKSCAN1* | intron | 0.150 | 0.038 | 1.27E-23 | 0.0011383 |
| 11:1219991:G:T | rs35705950 | *MUC5B* | upstream gene | 1.017 | -0.112 | < 1E-300 | 1.22E-08 |
| 13:112881427:C:T | rs12585036 | *ATP11A* | non coding transcript<br>exon | -0.121 | 0.063 | 2.35E-11 | 1.80E-06 |
| 16:276685:G:A | rs184954013 | *ARHGDIG* | intron | 0.640 | 0.358 | 1.30E-13 | 0.0078346 |
| 17:46126154:C:T | rs113120855 | *KANSL1* | intron | -0.216 | -0.084 | 8.92E-23 | 1.04E-08 |
| 19:4717660:A:G | rs12610495 | *DPP9* | missense | 0.109 | 0.108 | 2.95E-11 | 6.09E-18 |

**Figure 2.**
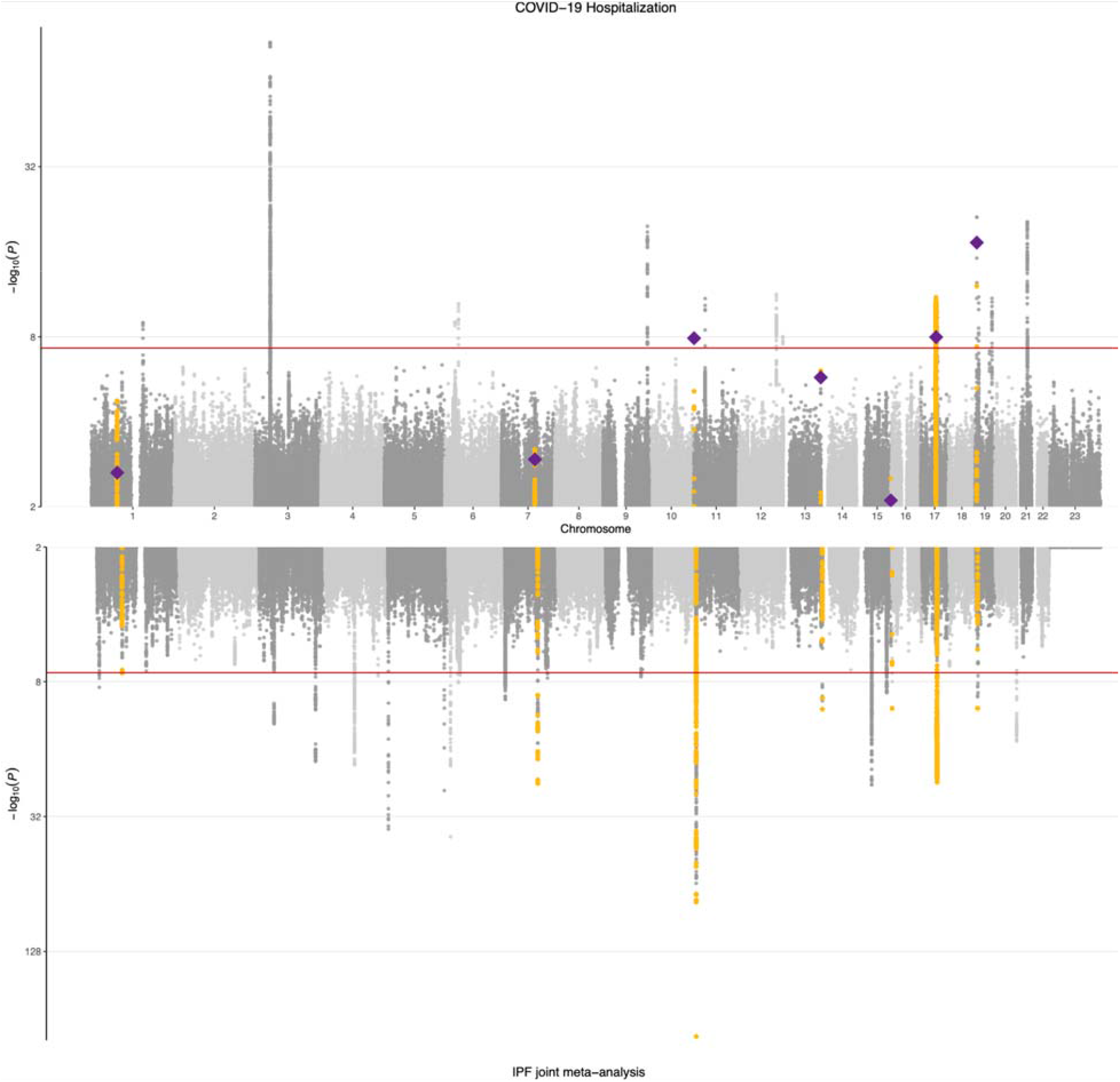
IPF meta-analysis and COVID-19 hospitalization results. COVID-19 hospitalization results are shown in the top panel and IPF joint meta-analysis results in the bottom panel. Genome-wide significant signals that reached FDR-adjusted p-value < 0.05 in COVID-19 hospitalization scan are highlighted in yellow. Index variants in IPF are plotted as diamonds in the COVID-19 results.

**Figure 3.**
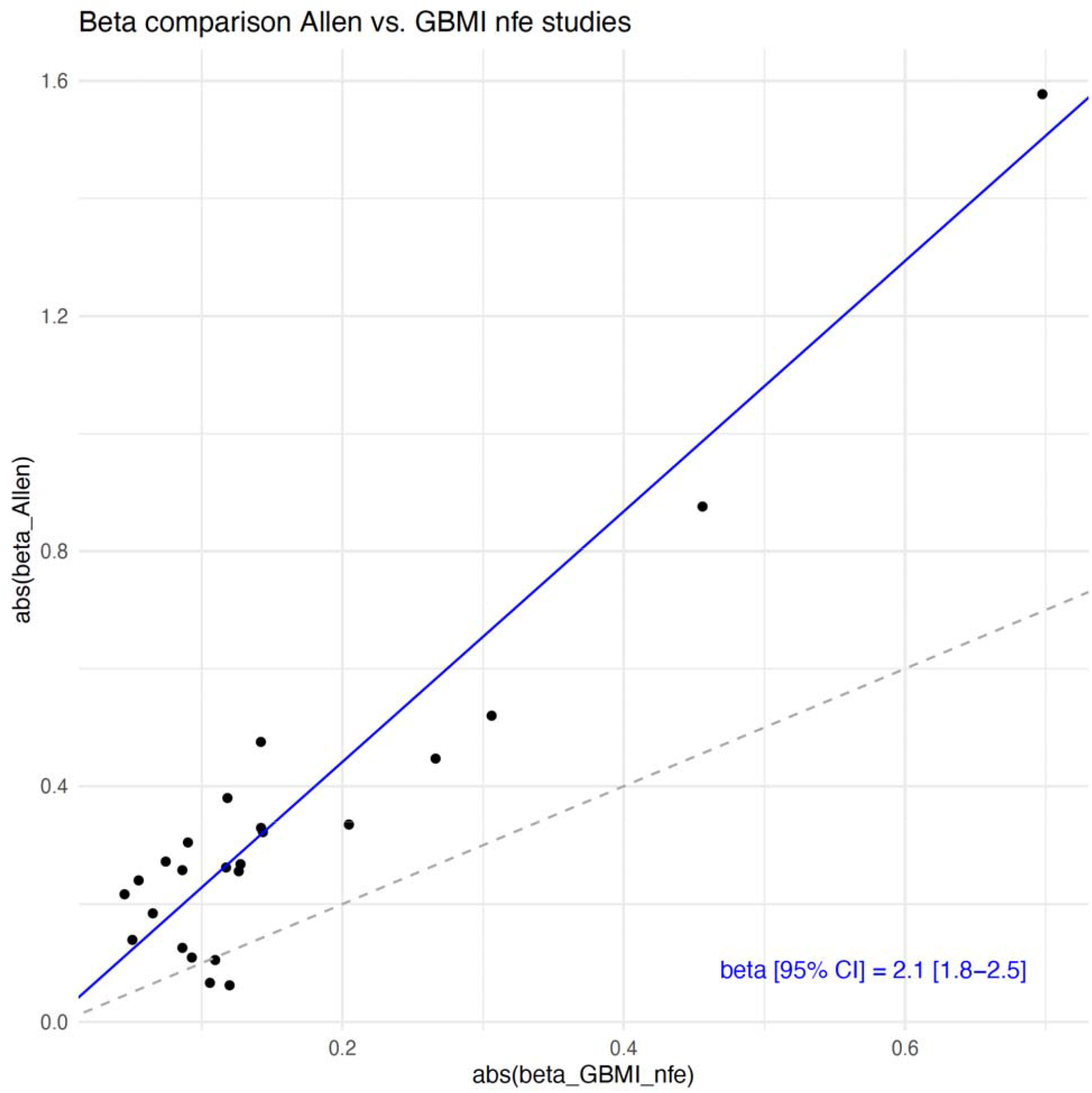
Effect size estimate comparison GBMI vs. latest IPF meta-analysis. Scatter plot of absolute value of latest IPF meta-analysis beta against absolute value of meta-analyzed GBMI NFE beta with inverse variance weighted linear regression line (weights from Allen et al. study) and accompanying slope estimate. The analysis was performed within the non-Finnish European ancestry and variants included were genome-wide significant in the joint meta-analysis.

Formal colocalization analysis was not possible as COVID-19 results have not been fine-mapped. Genetic correlation determined by LDSC^30^ between the traits was 0.31 (95% CI 0.15-0.47, p = 0.0001), in complete agreement with the previous estimate^14^ but with less uncertainty.

Pleiotropy of the IPF signals beyond COVID-19 was explored by colocalization analysis in FinnGen, pointing to shared signals between IPF and osteoporosis, cancers, and hypothyroidism (Supplementary Results).

### Sex-stratified analysis identifies heterogenic effects at *MUC5B*

Sex-stratified meta-analysis in the GBMI identified a 1.6-fold larger effect for the strongest IPF associated variant rs35705950 in the *MUC5B* locus in males (OR[95%CI] = 3.50[3.11-3.93], p = 8.5E-100) compared with females (OR[95%CI] = 2.20[1.91-2.52], p = 2.06E-29), Cochran’s Q p-value for heterogeneity = 3.58E-07. To investigate whether the difference in effects was due to confounding by case ascertainment differences across contributing biobanks, we assessed the effect of rs35705950 in males and females, noting a weaker effect in females across the biobanks (Figure S3, Table S7). MUC5B carrier status did not have an effect on the number of IPF deaths or lung transplants among IPF cases in FinnGen (Table S8).

The male only meta-analysis also identified two additional loci: 19q13.32 index variant rs71338787 in *EML2* and Xq28 index variant rs5945238 in *MPP1* (Table S9).

### Heterogeneity assessment points to large effect of sample ascertainment

As we observed heterogeneous effects across biobank and ancestry at nearly half of the IPF genome-wide significant loci (11 / 25, 44%, FDR-adjusted Cochran’s Q p-value < 0.05, mean heterogeneity index I^2^ = 0.62, Figure S4, Table S2), we explored whether there was a systematic difference between the effects observed in the latest IPF meta-analysis, involving carefully curated clinically defined IPF, and biobank defined IPF, generally coming from ICD-codes in electronic health records. Limiting to samples of non-Finnish European descent and genome-wide significant loci in the meta-analysis, the effect size estimates were 2.1-times larger in the clinical cohort compared to the meta-analyzed biobank studies (p = 1.6E-10, Figure 4). Per each biobank, the median beta-ratio (β_GBMI_/β_Allen_) varied from -0.04 to 0.62 (Figure S4).

To further study the effect of case ascertainment on effect size estimates, we divided FinnGen into three subsets based on diagnosis and original study cohort: a clinical IPF cohort (FinnishIPF^31^, n cases = 205), other IPF patients (n = 1,366), and non-IPF ILD patients (n = 1,624) and compared effect size estimates from these cohorts to those of the latest IPF meta-analysis. Again, effect size estimates were 0.9, 1.4, and 2.5-times larger in the latest IPF meta-analysis compared with the FinnishIPF, other IPF, and non-IPF ILD cohorts (Figure S6), providing further evidence that effect sizes in highly ascertained IPF patients are substantially higher compared with patients identified from biobanks.

The large effect of case ascertainment on effect size estimates was corroborated by meta-regression results, where case ascertainment explained most of the observed heterogeneity, while ancestry, by comparison, explained very little (mean R^2^ 55.8% and 6.2%, respectively, Supplementary Results).

## Discussion

We conducted the first multi-ancestry meta-analysis of IPF, increasing the number of IPF cases over 4-fold compared to the latest IPF meta-analysis. Incorporating 11,160 patients from six ancestries allowed us to identify seven novel loci associated with IPF susceptibility. One of the loci was only polymorphic in the East Asian population and two index variants were enriched within the meta-analysis over twofold in a non-European population compared to non-Finnish Europeans. Moreover, only one of the loci would have reached genome-wide significance had the analysis been restricted to European populations. Three of the identified loci, *GPR157, FKBP5*, and *RAPGEF2* have been previously associated with lung function measurements.

Fine-mapping in the Finnish population, enriched for deleterious low-frequency coding variants, suggested a predicted missense variant causal at the *KIF15* locus. In future studies, after further increases in non-European ancestry sample sizes, fine-mapping in multiple populations should be undertaken. Cross-ancestry fine-mapping, however, still has notable challenges to be resolved^32^.

COVID-19 severity and IPF share a notable proportion of their genetic background, as genetic correlation was estimated to be substantial (R_g_∼0.31), in line with previous estimates^14^ but with far less uncertainty. Of the 25 IPF index variants, seven reached the FDR-adjusted p-value of less than 5% in the COVID-19 hospitalization scan (out of which three were genome-wide significant: *CRHR1*, in addition to the previously reported *MUC5B* and *DPP9*). Effects at these loci were mostly in the same direction for the two traits, but variants in *MUC5B* and *ATP11A* showed opposite directions of effects. The strongest IPF risk locus *MUC5B* confers protection from severe COVID-19 and has been previously associated with improved survival in IPF patients^33^, although we observed no differences in the number of IPF-specific deaths or lung transplants across *MUC5B* carrier status among IPF cases in FinnGen. Acute exacerbations of IPF have poor prognosis, and present with Diffuse Alveolar Damage (DAD), which is also present in severe COVID-19^34^. Whether the mechanisms by which *MUC5B* confers protection from severe COVID-19 and may improve survival in IPF are shared, for example by protection from DAD, requires further research.

The increase in power conferred by the meta-analysis enabled subgroup and downstream analyses, allowing further examination of associations at both previous and novel loci. Interestingly, we observe a sex-stratified effect at *MUC5B*, the strongest known IPF risk factor, where the effect is 1.6 times larger in males compare to females. The sex-stratified effect may, however, arise from confounding by factors such as ascertainment and age distribution differences in the sexes and between-study heterogeneity in the context of differing male to female balance among the included studies. These confounders should be investigated in future studies before the observed difference can be claimed to represent a biological difference between the sexes.

The meta-analysis with contributing biobanks featuring a wide variety of sampling strategies enabled studying between-study heterogeneity, which revealed that case ascertainment has a large effect on IPF effect size estimates. Nearly a half of the genome-wide significant IPF loci expressed evidence of heterogeneity, which was mainly due to differences in case ascertainment – whether the patients were recruited from a hospital’s pulmonary clinic or identified from health registries. Effect size estimates were around two-fold for clinical IPF-cohorts compared to patients recruited from biobanks, suggesting misclassification of IPF in biobanks. For genome-wide association studies, however, the substantially larger number of patients available from biobanks benefits discovery even given the attenuated effect size estimates.

Limitations of the study include between-study heterogeneity, pointing to differing ascertainment between studies. However, this has limited impact on our novel findings, as only one of the novel loci showed evidence of heterogeneity (*DNAJB4*). Second, the PheCode-based IPF case definition was marginally more inclusive than the conventional definition, increasing the risk of misclassification in biobanks. Third, even though samples representing four non-European ancestries were included, the sample was still dominated by participants of European ancestry. Finally, fine-mapping was successful for only a minority of the loci.

We present the first multi-ancestry meta-analysis of IPF with an over 4-fold increase in cases compared to the latest IPF meta-analysis. Multiple novel loci are discovered, the vast majority of which are driven by non-European populations and many of which have been linked to lung traits. We confirm and further describe the notable overlap of genetic determinants of IPF and severe COVID-19, calling for functional research. We describe a sex-dependent effect at the strongest IPF risk factor, the *MUC5B* locus, and demonstrate a two-fold difference in effect size estimates derived from clinical cohorts as opposed to biobanks. To conclude, leveraging global cross-population analysis further elucidates the genetic background of IPF by both revealing novel loci and providing increased resolution into previously identified ones.

## Material and Methods

### Study cohorts

13 biobanks in Europe, Asia, and USA encompassing 6 ancestries contributed to the Global Biobank Meta-analysis Initiative (GBMI) IPF meta-analysis, totaling 8,492 cases and 1,355,819 controls (Table 1, Table S1). Sample recruitment strategies differed between the biobanks (Table S1). In FinnGen sex-stratified analyses were conducted in release 5 with 378 and 110524, and 650 and 86462 female and male IPF cases and controls, respectively. The three clinical IPF cohorts of the latest IPF meta-analysis (Chicago, Colorado and UK), totaling 2,668 cases and 8,591 controls, all of European ancestry, are described elsewhere^2^. The two cohorts used for replication were also used for replication in the latest meta-analysis and are described elsewhere^2^. There was sample overlap of n = 3,366 controls between the GBMI meta-analysis and the three clinical IPF cohorts of the latest IPF meta-analysis, but there was no IPF case overlap.

### Phenotype definition and quality control

The GBMI phenotypic and genotypic quality control are described elsewhere^35^. Analysis was mainly performed for PheCode 502, constructed from health data available from each biobank (Table S10, phenotype definitions used in each biobank are in Table S11). IPF cases for the PheCode 502 were determined using the following International Classification of Diseases (ICD)-codes: ICD-9: 515, 515.0, ICD-10: J84.1, J84.10, J84.17, J84.8, J84.89. In the latest IPF meta-analysis, case definition was based on American Thoracic Society and European Respiratory Society guidelines, and quality control steps are described elsewhere ^2^.

### Meta-analysis

For GBMI, GWASs stratified by ancestry and sex were conducted in each biobank after standard sample-level and variant-level quality control and fixed-effect meta-analyses based on inverse-variance weighting were performed for all biobanks across all ancestries, and all biobanks by sex, detailed description elsewhere^35^. The GBMI meta-analysis was performed for both sexes, and for males and females separately. Meta-analysis of the GBMI meta-analysis and the latest IPF meta-analysis, hereafter referred to as the joint meta-analysis, was likewise performed using the inverse-variance weighted fixed effects model in R (version 4.1). Prior to meta-analysis, the summary statistics of the latest IPF meta-analysis were lifted over from GRCh37 to GRCh38 using UCSC liftOver (https://genome.ucsc.edu/cgi-bin/hgLiftOver). Liftover results were verified by comparing the results to LiftOver results from Picard (Supplementary Results). All variants are reported based on the human genome reference sequence GRCh38. The LD score regression intercept^17^ was used to explore possible confounding by e.g. population stratification. Only variants that were genome-wide significant after joint meta-analysis (of GBMI and Allen) were considered in downstream analysis. Genome-wide significant loci were determined by taking a 1 Mb region around each genome-wide significant variant and merging overlapping regions. The HLA region on chromosome 6 (GRCh38 chr6:28,510,120-33,480,577) was considered one locus.

### Fine-mapping

Fine-mapping was performed for the IPF GWAS in FinnGen release 7 using the “Sum of Single Effects” (SuSie) model ^36,37^ for each genome-wide significant locus. Fine-mapping regions were defined by taking a 3 Mb window around each index variant and merging overlapping regions. 95% credible sets (encompassing at least 95% of the probability of including the causal variant) were analyzed and the probability of variant causality was evaluated using the posterior inclusion probability (PIP). Conditional analysis in FinnGen was run using REGENIE ^38^ with dosages of the variant conditioned on as covariates alongside other covariates (age, sex, ten first principal components, and batch). Logistic regression with Firth correction exploring individual causal signals was performed in FinnGen using the “logistf” R package^39^.

### Phenome-wide lookup

To assess the shared effects of potentially novel loci, we considered phenotypes in Open Targets Genetics (OTG) obtained from the GWAS catalog. Linkage disequilibrium (LD) between variants was assessed using the LD pair tool^40^ (https://ldlink.nci.nih.gov/), restricting the analysis to the 1000 Genomes Project non-Finnish European sub-populations.

### Colocalization

Colocalization analysis was performed in FinnGen release 7. Colocalization analysis was based on assessing agreement of the fine-mapped credible sets across two traits. Agreement was measured by causal posterior agreement (CLPA), calculated as the sum of minimum PIP between the two traits per variant in overlapping credible sets.

### Longitudinal analysis

Descriptive longitudinal analyses were based on Aalen-Johansen estimates of cumulative incidence to account for competing risk of other causes of death. Illustrations are graphically smoothed to respect the privacy of study participants.

### Heterogeneity evaluation

Heterogeneity of effect sizes across studies and between sexes was evaluated at each variant using Cochran’s Q p-value and the heterogeneity index. To study the contribution of different sample recruitment strategies on heterogeneity, effect size estimates of selected studies for genome-wide significant IPF loci were compared and an inverse variance weighted linear regression line was fitted. To evaluate the extent to which sample recruitment and ancestry contributed to heterogeneity, we used meta-regression ^41^ using the “meta” R package^42^.

## Data Availability

Full summary statistics and code are available per request.

## Acknowledgements

We would like to thank Jaakko Kaprio from the University of Helsinki for comments on the manuscript. This research used the SPECTRE High Performance Computing Facility at the University of Leicester.

This work was supported by the Doctoral Programme in Population Health, University of Helsinki [to JJP]; and The Finnish Medical Foundation [to JJP]; RJA is an Action for Pulmonary Fibrosis Mike Bray Research Fellow, NHLBI [to JO]; Wellcome Trust to [BGG]; Spanish Ministry of Science and Innovation and Instituto de Salud Carlos III, co-financed by by the European Regional Development Fund (ERDF) “A Way of Making Europe” from the European Union (EU) to [CF]; Cabildo Insular de Tenerife to [CF]; NIH [to IN], Medical Research Council -Project Grant [to RGJ]; NIHR Research Professorship [to RGJ]; GSK/British Lung Foundation Chair in Respiratory Research (C17-1) [to LVW]; Research Foundation of the Pulmonary Diseases HES [to RK]; Jalmari and Rauha Ahokas Foundation [to RK]; the Research Foundation of North Finland, Oulu, Finland and a state subsidy of Oulu University Hospital [to RK]; HUS State Research Funding [to MM]. The work of the contributing biobanks for GBMI was supported by numerous grants from governmental and charitable bodies, see Supplementary Material. The research was partially supported by the National Institute for Health Research (NIHR) Leicester Biomedical Research Centre; the views expressed are those of the author(s) and not necessarily those of the National Health Service (NHS), the NIHR or the Department of Health.

## Data and Code availability

Full summary statistics and code are available per request.

