## Supplementary Material for "Leveraging global multi-ancestry meta-analysis in the study of Idiopathic Pulmonary Fibrosis genetics"

### Supplementary Results

#### Verification of LiftOver results

Of the 10,790,934 variants lifted over by UCSC liftOver 10,777,976 (99.9%) overlapped with LiftOver results from Picard, indicating high concordance in the results of the two methods.

#### Fine-mapping of the 11p15.5 locus

Fine-mapping indicated two independent signals at 11p15.5; the well-established *MUC5B* and an independent signal downstream to *MOB2.* Causality of the *MOB2* downstream variant (rs546531844, AF = 0.29%, 17-times enriched in Finns compared NFEE, gnomAD v2.1.1) was corroborated by two different fine-mapping methods.

The "Sum of Single Effects" (SuSie) model[1] suggested two 95% credible sets at the 11p15.5 locus. One of the credible sets included the *MUC5B* upstream gene variant rs35705950G>T (posterior inclusion probability (PIP) = 1) and the other included two variants: a *MOB2* downstream gene variant rs546531844G>A (PIP = 0.92) and a *MUC6* missense variant rs148815783C>T (PIP = 0.072).

Another fine-mapping method, FINEMAP [2,3], suggested four 95% credible sets (prob = 0.55) at the locus. Three of the four credible sets consisted of one variant. Both the *MUC5B* upstream gene variant rs35705950G>T and the *MOB2* downstream gene variant rs546531844G>A constructed their own one-variant credible sets (PIP = 1 and PIP = 0.96, respectively).

After conditioning the REGENIE GWAS on the *MUC5B* variant rs35705950G>T the *MOB2* downstream gene variant rs546531844G>A association was no longer genome-wide significant and the effect size estimate was notably decreased (original beta = 1.35, original p = 1.33E-37, conditioned beta = 0.53, conditioned p = 1.41E-7).

Imputation accuracy of the *MOB2* variant was not optimal (INFO = 0.85) and LD between the *MOB2* downstream variant and the *MUC5B* lead variant was low but not inexistent ($r^2$ = 0.0721, D' = 0.7658). Thus, the signal at *MOB2* remains to be confirmed.

#### FinnGen colocalization analysis

Causal posterior agreement (CLPA, measuring the agreement between credible sets over two traits when overlapping credible sets are assessed) was used to define colocalization beyond COVID-19 in a two-trait model. At 16p13.3 an intronic *ARHGDIG* variant (rs184954013, AF in Finns ~2.3%, AF in NFE ~0.025) colocalized with osteoporotic fracture signal (CLPA=0.74) and also with any form of osteoporosis (CLPA=0.58). As we have reported prior to this study[14], multiple malignancy related colocalization signals were also detected (Table S11).

**Dissecting contributors to heterogeneity by metaregression**

After adjusting for ancestry, all 11 loci expressed evidence of remaining heterogeneity with a mean R-squared of only 6.2 for ancestry. To study the effect of case ascertainment, we compared the effect estimates of two clinical cohorts, the latest IPF meta-analysis and a subcohort of FinnGen (FinnishIPF, n cases = 205), to the estimates of the 13 GBMI biobanks (excluding the clinical subcohort from FinnGen). Having divided FinnGen into two sub-cohorts, 10 of the 25 loci expressed evidence of heterogeneity. However, after accounting for case ascertainment status (whether the studies were based on clinical cohorts or not), only 4 of these 10 loci expressed evidence of remaining heterogeneity. The mean R-squared in the 10 loci was 55.8 for clinical cohort status.

### Supplementary Figures

**Fig S1.** Locus zoom plots for all 25 genome-wide significant loci.

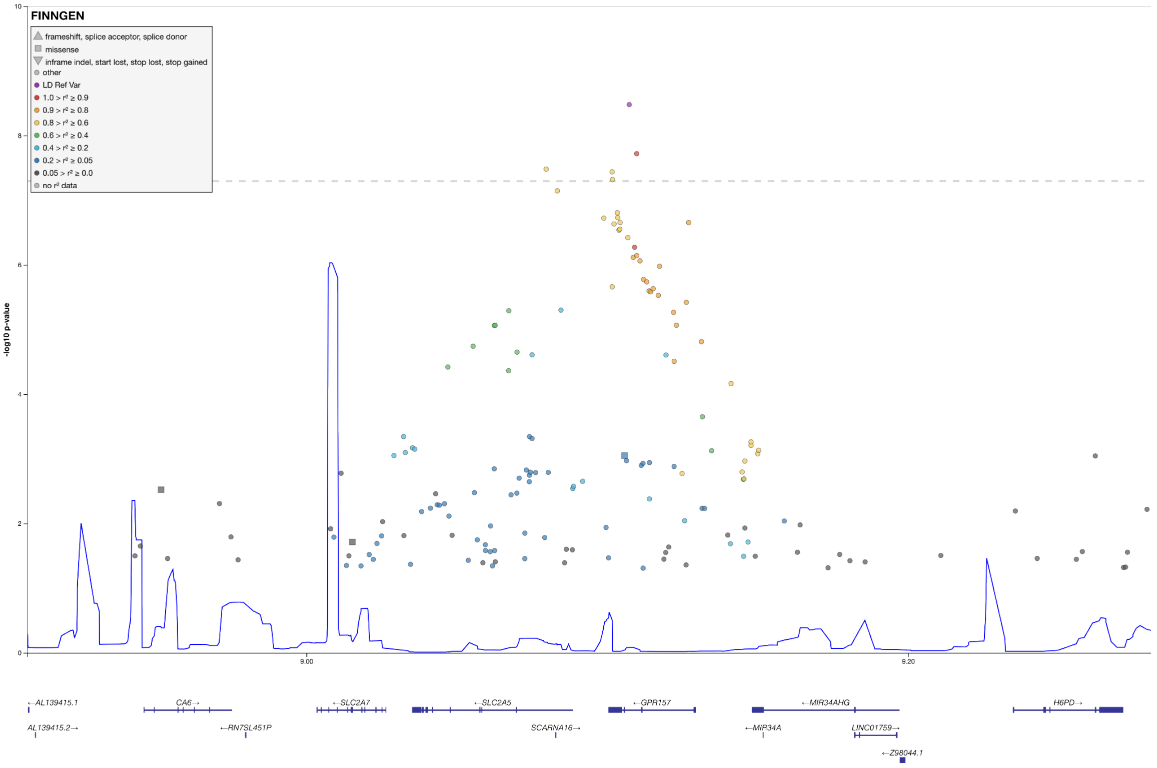


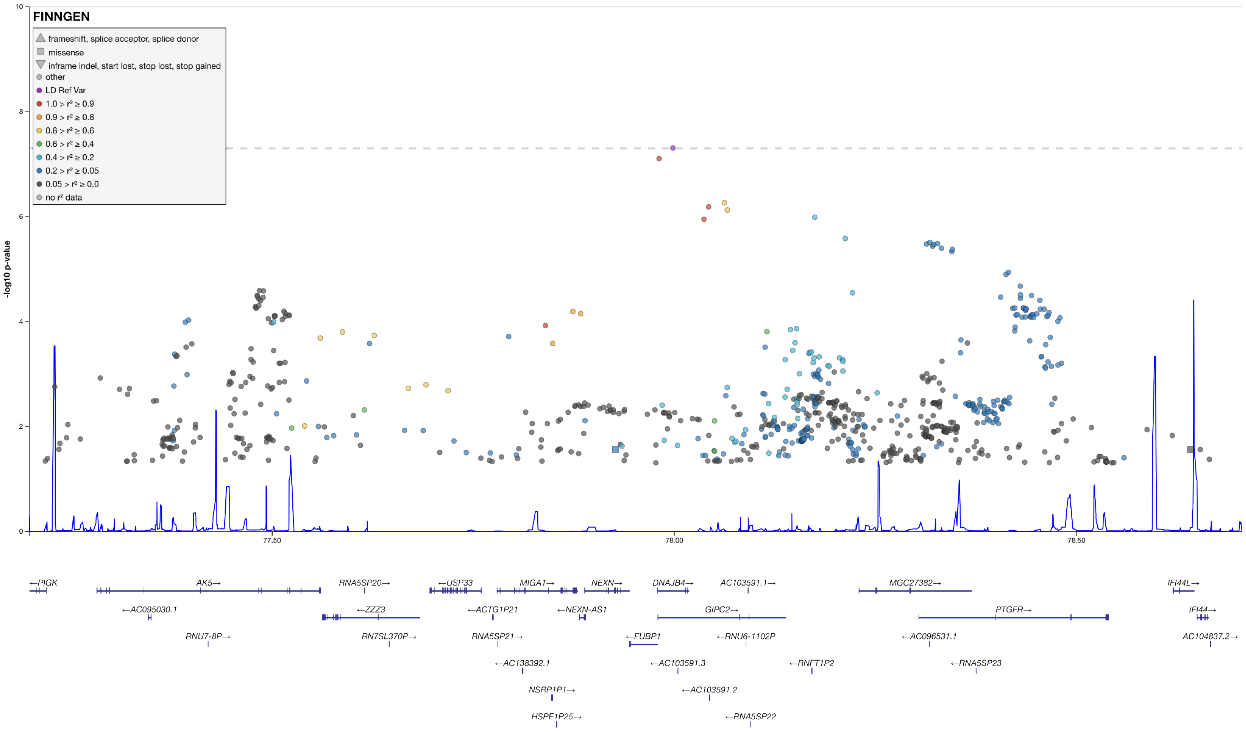

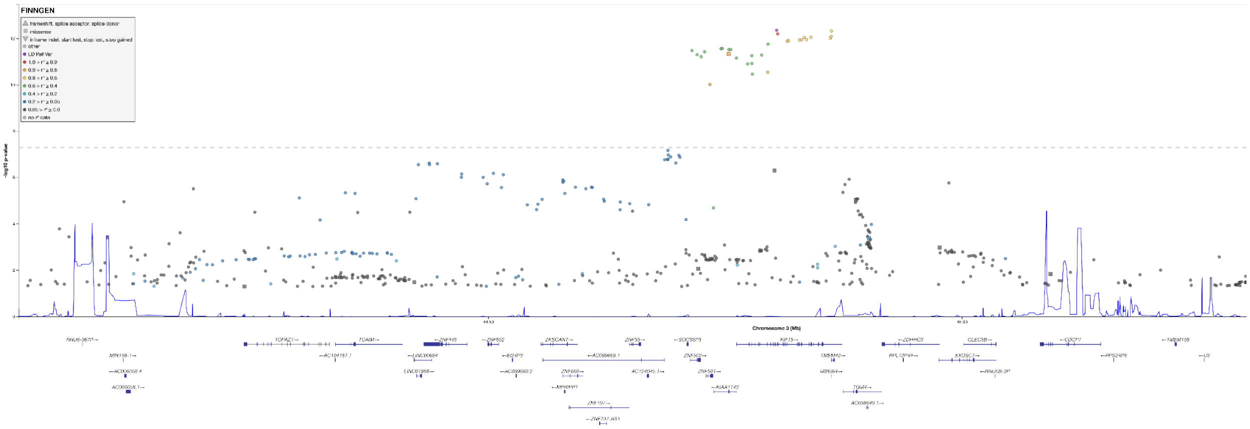
FINNGEN
-log10 p-value
Chromosome 3 (Mb)

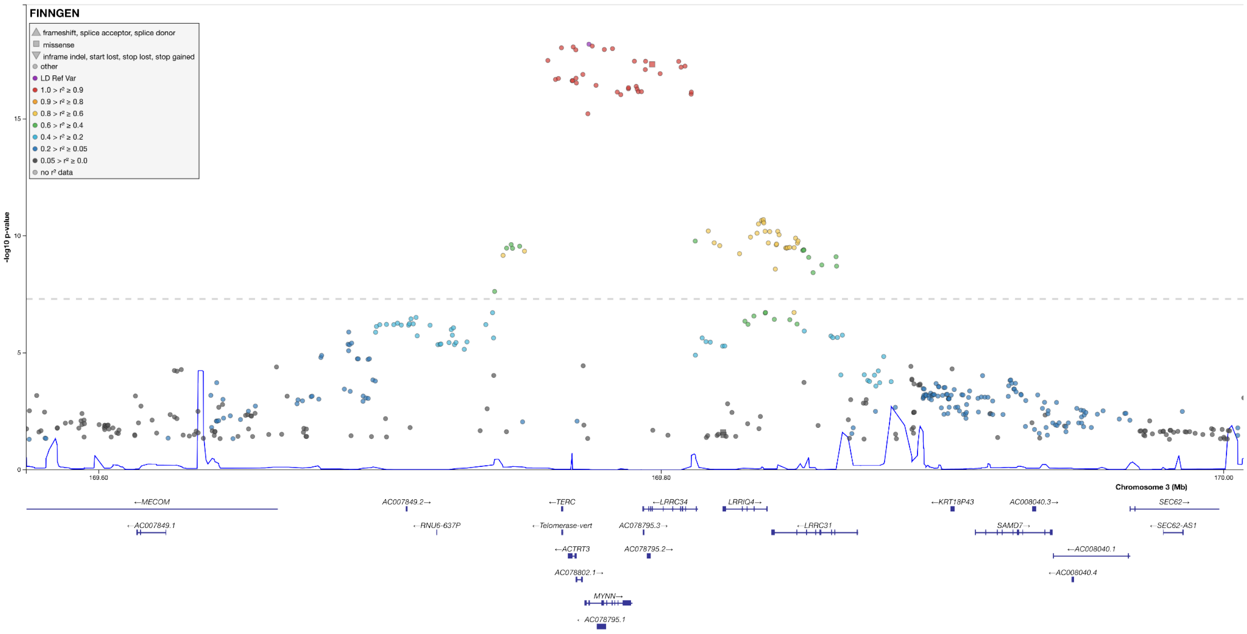
FINNGEN
frameshift, splice acceptor, splice donor
missense
inframe indel, start lost, stop lost, stop gained
other
LD Ref Var
1.0 > r² ≥ 0.9
0.9 > r² ≥ 0.8
0.8 > r² ≥ 0.6
0.6 > r² ≥ 0.4
0.4 > r² ≥ 0.2
0.2 > r² ≥ 0.05
0.05 > r² ≥ 0.0
no r² data
-log10 p-value
15
10
5
0
169.60
169.80
170.00
Chromosome 3 (Mb)
←MECOM
AC007849.2→
←TERC
←LRRC34
LRRIQ4→
←KRT18P43
AC008040.3→
SEC62→
←AC007849.1
←RNU6-637P
←Telomerase-vert
AC078795.3→
←LRRC31
SAMD7→
←SEC62-AS1
←ACTRT3
AC078795.2→
←AC008040.1
AC078802.1→
←AC008040.4
MYNN→
←AC078795.1

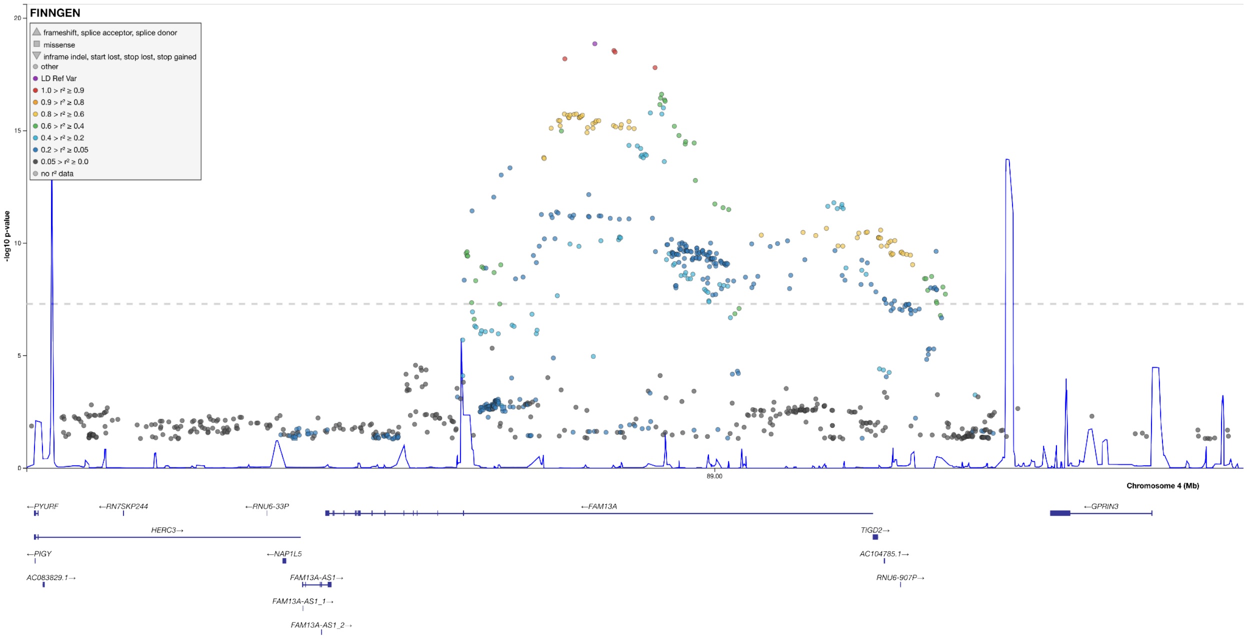
FINNGEN
frameshift, splice acceptor, splice donor
missense
inframe indel, start lost, stop lost, stop gained
other
LD Ref Var
1.0 > r² ≥ 0.9
0.9 > r² ≥ 0.8
0.8 > r² ≥ 0.6
0.6 > r² ≥ 0.4
0.4 > r² ≥ 0.2
0.2 > r² ≥ 0.05
0.05 > r² ≥ 0.0
no r² data
-log10 p-value
20
15
10
5
0
89.00
Chromosome 4 (Mb)
←PYURF
←RN7SKP244
←RNU6-33P
←FAM13A
←GPRIN3
HERC3→
TIGD2→
←PIGY
←NAP1L5
AC104785.1→
AC083829.1→
FAM13A-AS1→
RNU6-907P→
FAM13A-AS1_1→
FAM13A-AS1_2→

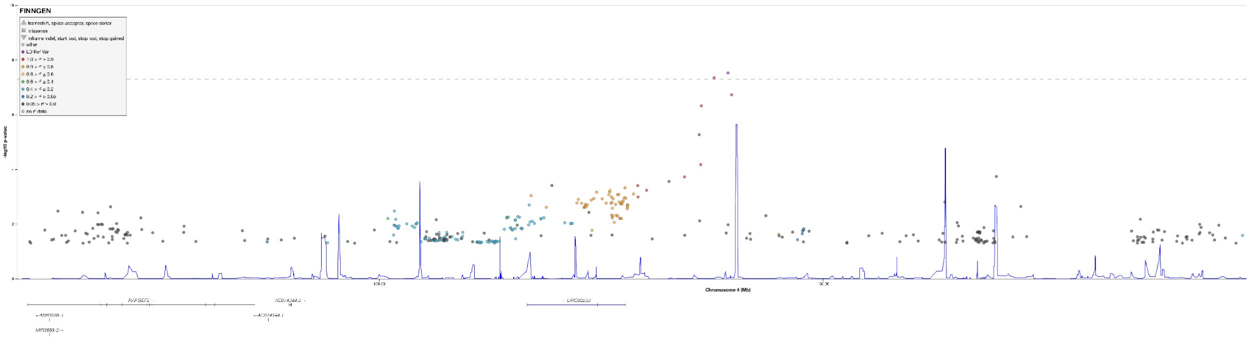
FINNGEN
-log10 p-value
Chromosome 4 (Mb)

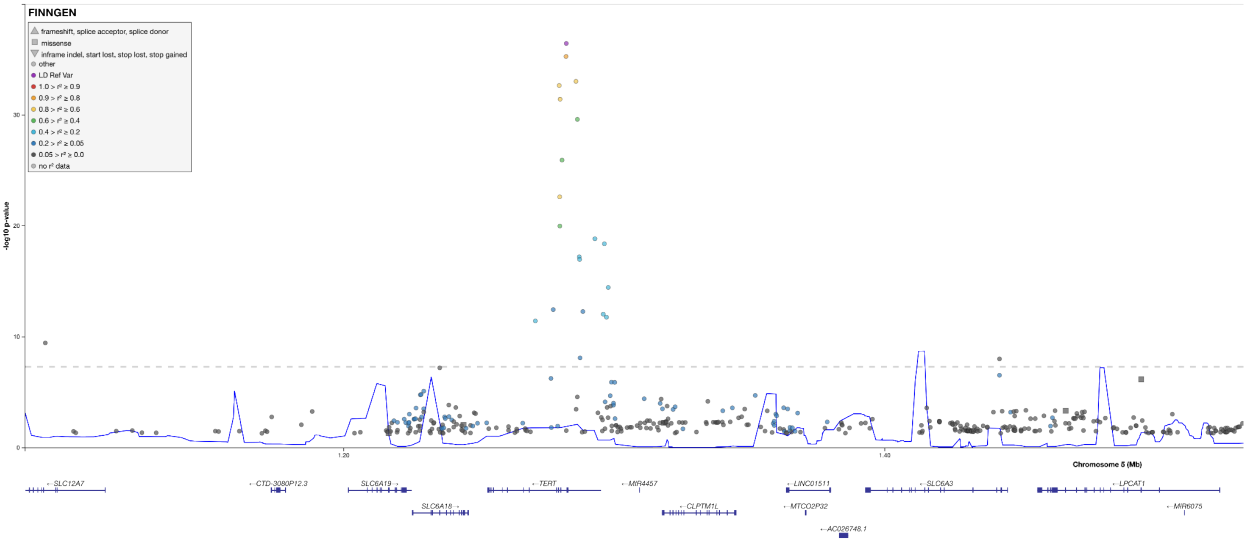
FINNGEN
frameshift, splice acceptor, splice donor
missense
inframe indel, start lost, stop lost, stop gained
other
LD Ref Var
1.0 > r² ≥ 0.9
0.9 > r² ≥ 0.8
0.8 > r² ≥ 0.6
0.6 > r² ≥ 0.4
0.4 > r² ≥ 0.2
0.2 > r² ≥ 0.05
0.05 > r² ≥ 0.0
no r² data
-log10 p-value
30
20
10
0
1.20
1.40
Chromosome 5 (Mb)
←SLC12A7
←CTD-3080P12.3
SLC6A19→
←TERT
←MIR4457
←LINC01511
←SLC6A3
←LPCAT1
SLC6A18→
←CLPTM1L
←MTCO2P32
←MIR6075
←AC026748.1

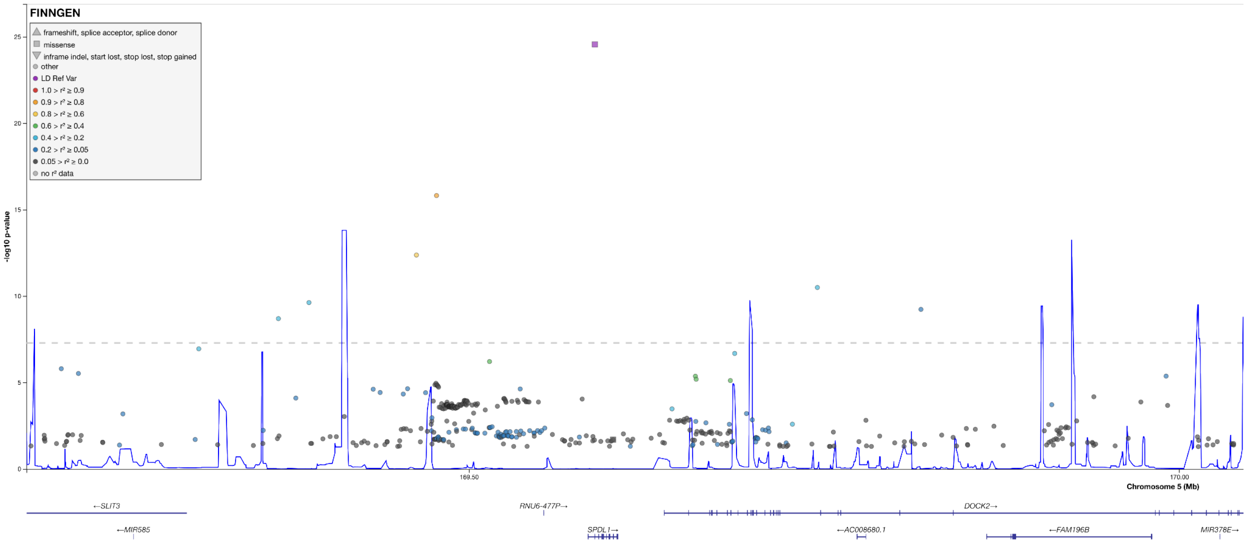
FINNGEN
frameshift, splice acceptor, splice donor
missense
inframe indel, start lost, stop lost, stop gained
other
LD Ref Var
1.0 > r² ≥ 0.9
0.9 > r² ≥ 0.8
0.8 > r² ≥ 0.6
0.6 > r² ≥ 0.4
0.4 > r² ≥ 0.2
0.2 > r² ≥ 0.05
0.05 > r² ≥ 0.0
no r² data
-log10 p-value
25
20
15
10
5
0
169.50
170.00
Chromosome 5 (Mb)
←SLIT3
RNU6-477P→
DOCK2→
←MIR585
SPDL1→
←AC008680.1
←FAM196B
MIR378E→

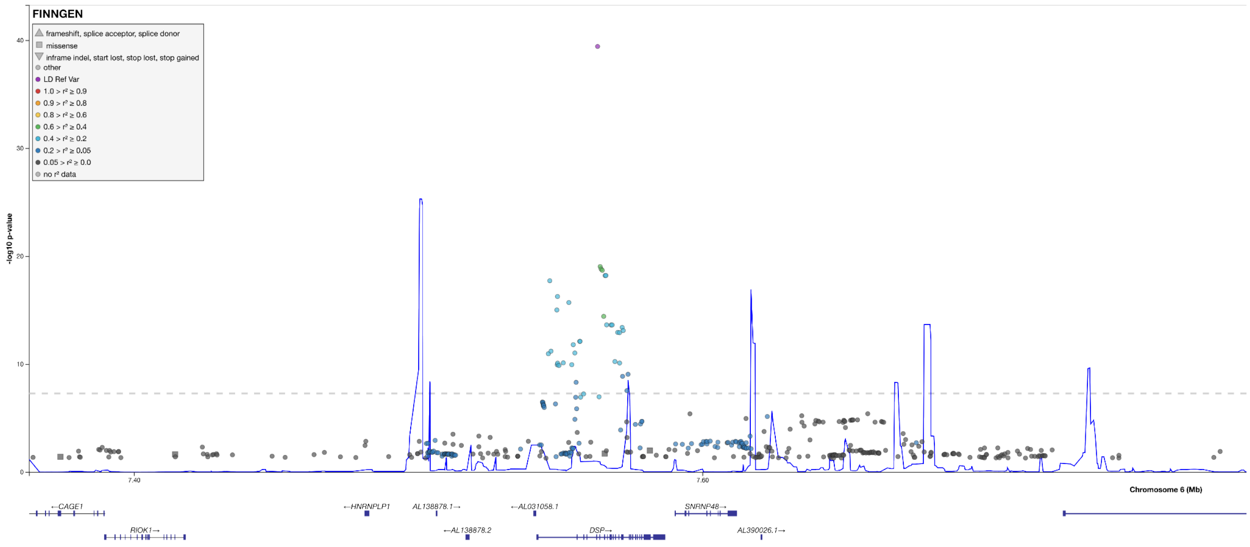
FINNGEN
frameshift, splice acceptor, splice donor
missense
inframe indel, start lost, stop lost, stop gained
other
LD Ref Var
1.0 > r² ≥ 0.9
0.9 > r² ≥ 0.8
0.8 > r² ≥ 0.6
0.6 > r² ≥ 0.4
0.4 > r² ≥ 0.2
0.2 > r² ≥ 0.05
0.05 > r² ≥ 0.0
no r² data
-log10 p-value
40
30
20
10
0
7.40
7.60
Chromosome 6 (Mb)
←CAGE1
RIOK1→
←HNRNPLP1
AL138878.1→
←AL138878.2
←AL031058.1
DSP→
SNRNP48→
AL390026.1→

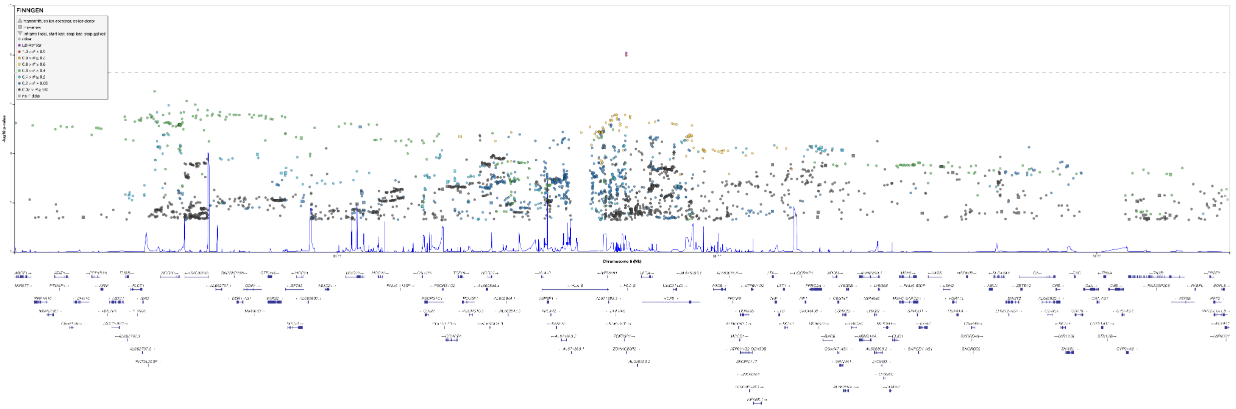
FINNGEN

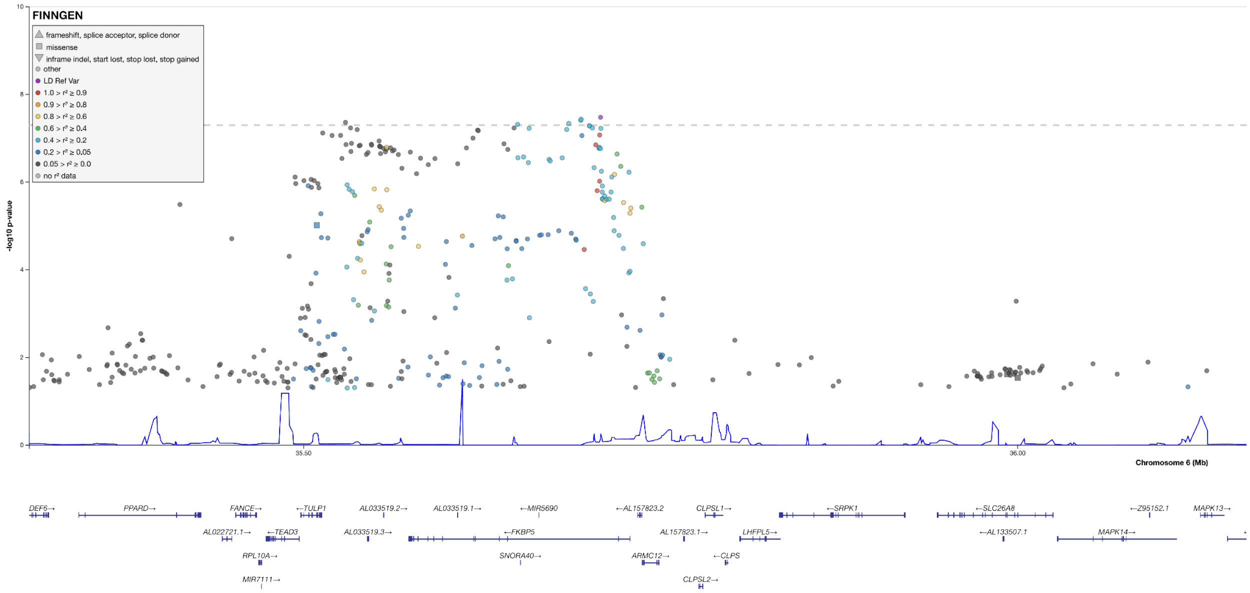
FINNGEN
frameshift, splice acceptor, splice donor
missense
inframe indel, start lost, stop lost, stop gained
other
LD Ref Var
1.0 > r² ≥ 0.9
0.9 > r² ≥ 0.8
0.8 > r² ≥ 0.6
0.6 > r² ≥ 0.4
0.4 > r² ≥ 0.2
0.2 > r² ≥ 0.05
0.05 > r² ≥ 0.0
no r² data
-log10 p-value
10
8
6
4
2
0
35.50
36.00
Chromosome 6 (Mb)
DEF6→
PPARD→
FANCE→
←TULP1
AL033519.2→
AL033519.1→
←MIR5690
←AL157823.2
CLPSL1→
←SRPK1
←SLC26A8
←Z95152.1
MAPK13→
AL022721.1→
←TEAD3
AL033519.3→
←FKBP5
AL157823.1→
LHFPL5→
←AL133507.1
MAPK14→
RPL10A→
SNORA40→
ARMC12→
←CLPS
MIR7111→
CLPSL2→

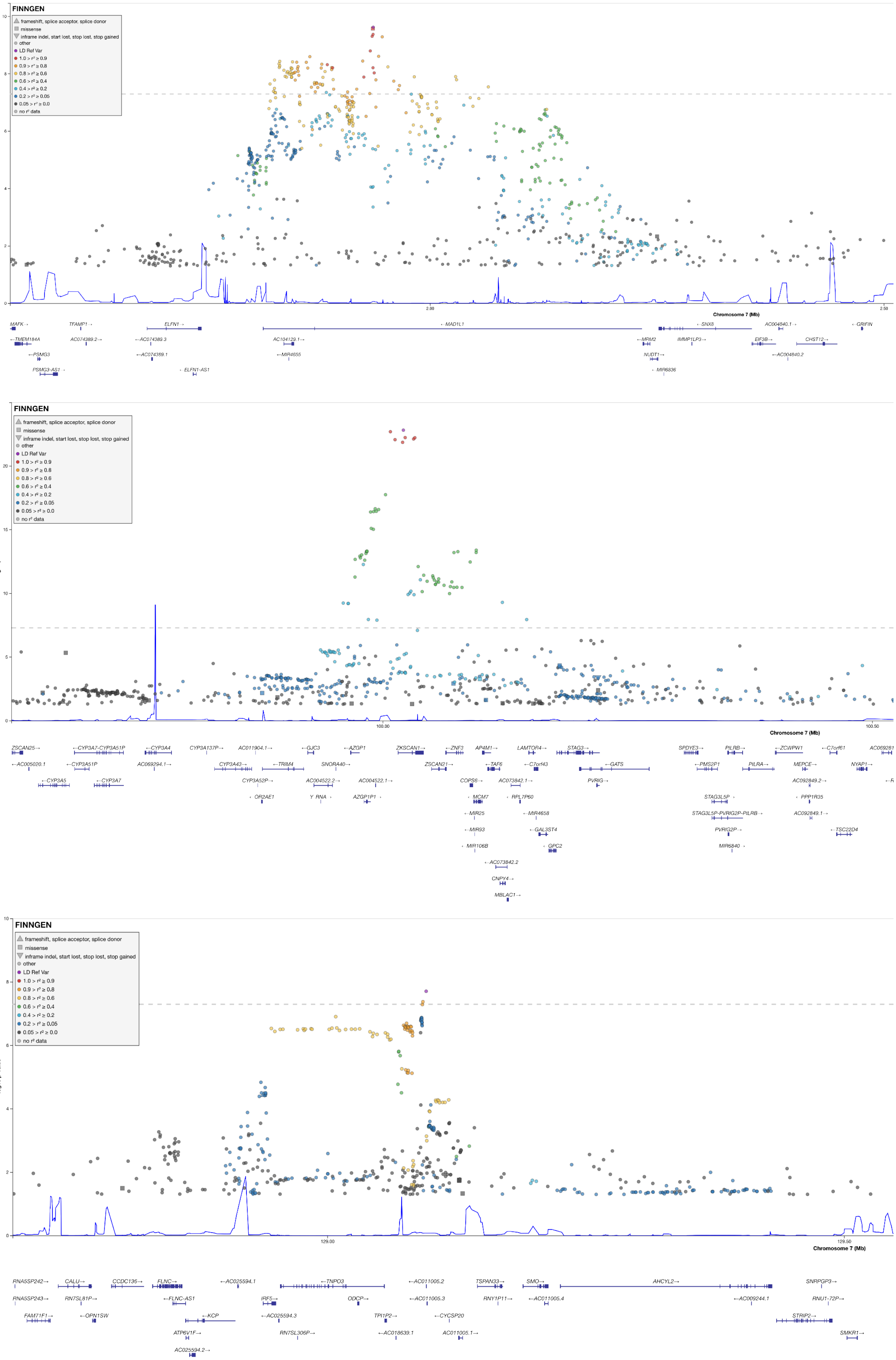
FINNGEN
frameshift, splice acceptor, splice donor
missense
inframe indel, start lost, stop lost, stop gained
other
LD Ref Var
1.0 > r² ≥ 0.9
0.9 > r² ≥ 0.8
0.8 > r² ≥ 0.6
0.6 > r² ≥ 0.4
0.4 > r² ≥ 0.2
0.2 > r² ≥ 0.05
0.05 > r² ≥ 0.0
no r² data
-log10 p-value
Chromosome 7 (Mb)
2.00
2.50
MAFK
TFAMP1
ELFN1
MAD1L1
SNX8
AC004840.1
GRIFIN
TMEM184A
AC074389.2
AC074389.3
AC104129.1
MRM2
IMMP1LP3
EIF3B
CHST12
PSMG3
AC074389.1
MIR4655
NUDT1
AC004840.2
PSMG3-AS1
ELFN1-AS1
MIR6836
FINNGEN
100.00
100.50
Chromosome 7 (Mb)
ZSCAN25
CYP3A7-CYP3A51P
CYP3A4
CYP3A137P
AC011904.1
GJC3
AZGP1
ZKSCAN1
ZNF3
AP4M1
LAMTOR4
STAG3
SPDYE3
PILRB
ZCWPW1
C7orf61
AC005020.1
CYP3A51P
AC069294.1
CYP3A43
TRIM4
SNORA40
ZSCAN21
TAF6
C7orf43
GATS
PMS2P1
PILRA
MEPCE
NYAP1
CYP3A5
CYP3A7
CYP3A52P
AC004522.2
AC004522.1
COPS6
AC073842.1
PVRIG
AC092849.2
OR2AE1
Y_RNA
AZGP1P1
MCM7
RPL7P60
STAG3L5P
PPP1R35
MIR25
MIR4658
STAG3L5P-PVRIG2P-PILRB
AC092849.1
MIR93
GAL3ST4
PVRIG2P
TSC22D4
MIR106B
GPC2
MIR6840
AC073842.2
CNPY4
MBLAC1
FINNGEN
-log10 p-value
129.00
129.50
Chromosome 7 (Mb)
RNA5SP242
CALU
CCDC136
FLNC
AC025594.1
TNPO3
AC011005.2
TSPAN33
SMO
AHCYL2
SNRPGP3
RNA5SP243
RN7SL81P
FLNC-AS1
IRF5
ODCP
AC011005.3
RNY1P11
AC011005.4
AC009244.1
RNU1-72P
FAM71F1
OPN1SW
KCP
AC025594.3
TPI1P2
CYCSP20
STRIP2
ATP6V1F
RN7SL306P
AC018639.1
AC011005.1
SMKR1
AC025594.2

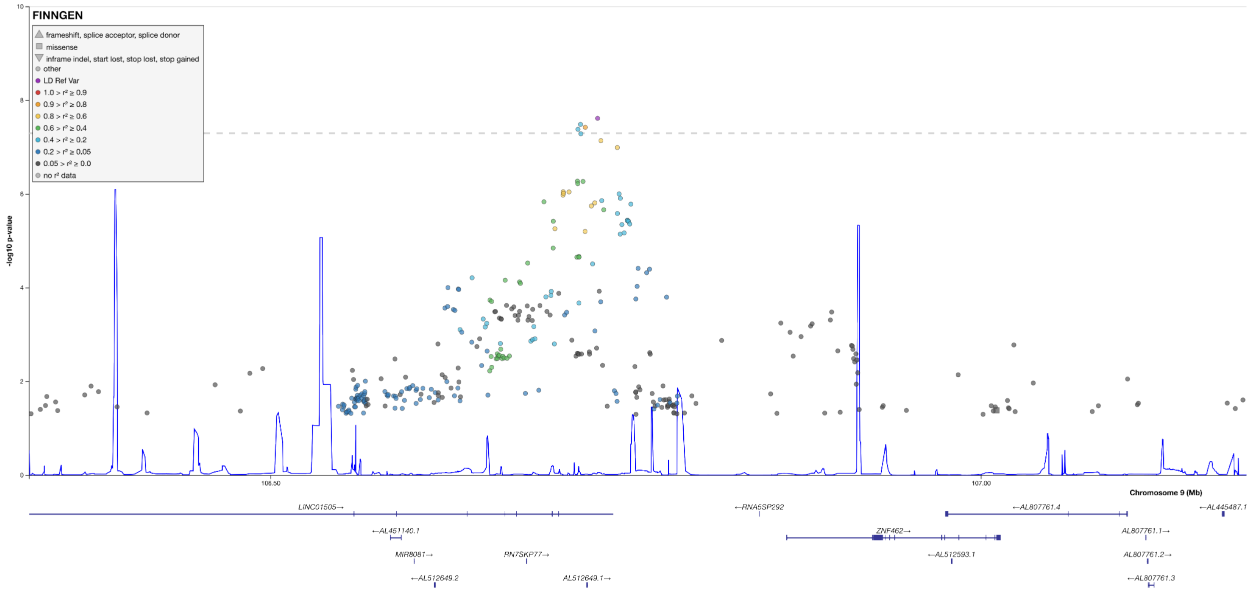
FINNGEN
frameshift, splice acceptor, splice donor
missense
inframe indel, start lost, stop lost, stop gained
other
LD Ref Var
1.0 > r² ≥ 0.9
0.9 > r² ≥ 0.8
0.8 > r² ≥ 0.6
0.6 > r² ≥ 0.4
0.4 > r² ≥ 0.2
0.2 > r² ≥ 0.05
0.05 > r² ≥ 0.0
no r² data
-log10 p-value
106.50
107.00
Chromosome 9 (Mb)
LINC01505→
←RNA5SP292
←AL807761.4
←AL445487.1
←AL451140.1
ZNF462→
AL807761.1→
MIR8081→
RN7SKP77→
←AL512593.1
AL807761.2→
←AL512649.2
AL512649.1→
←AL807761.3

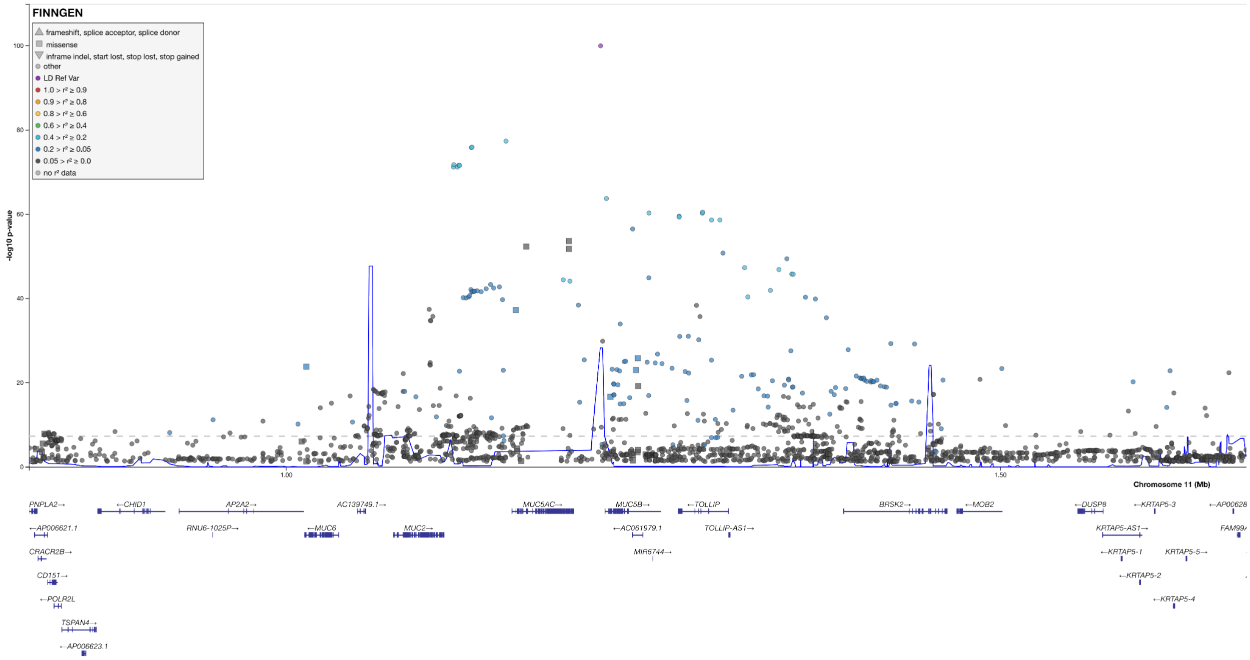
FINNGEN
frameshift, splice acceptor, splice donor
missense
inframe indel, start lost, stop lost, stop gained
other
LD Ref Var
1.0 > r² ≥ 0.9
0.9 > r² ≥ 0.8
0.8 > r² ≥ 0.6
0.6 > r² ≥ 0.4
0.4 > r² ≥ 0.2
0.2 > r² ≥ 0.05
0.05 > r² ≥ 0.0
no r² data
-log10 p-value
1.00
1.50
Chromosome 11 (Mb)
PNPLA2→
←CHID1
AP2A2→
AC139749.1→
MUC5AC→
MUC5B→
←TOLLIP
BRSK2→
←MOB2
←DUSP8
←KRTAP5-3
←AP006621.1
RNU6-1025P→
←MUC6
MUC2→
←AC061979.1
TOLLIP-AS1→
KRTAP5-AS1→
CRACR2B→
MIR6744→
←KRTAP5-1
KRTAP5-5→
CD151→
←KRTAP5-2
←POLR2L
←KRTAP5-4
TSPAN4→
←AP006623.1

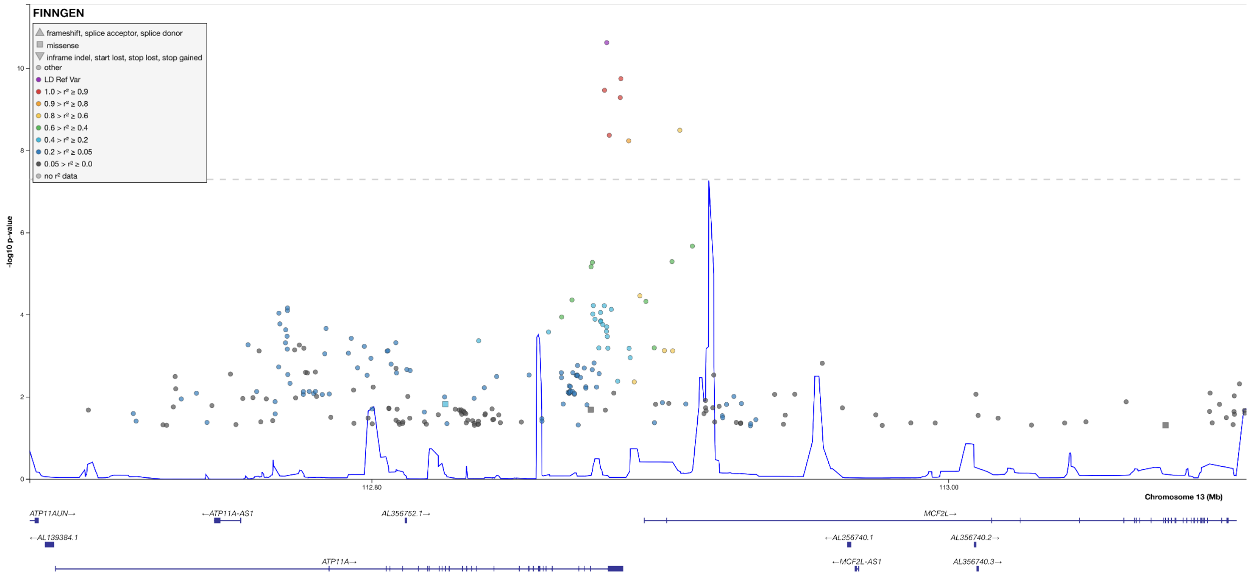
FINNGEN
frameshift, splice acceptor, splice donor
missense
inframe indel, start lost, stop lost, stop gained
other
LD Ref Var
1.0 > r² ≥ 0.9
0.9 > r² ≥ 0.8
0.8 > r² ≥ 0.6
0.6 > r² ≥ 0.4
0.4 > r² ≥ 0.2
0.2 > r² ≥ 0.05
0.05 > r² ≥ 0.0
no r² data
-log10 p-value
112.80
113.00
Chromosome 13 (Mb)
ATP11AUN→
←ATP11A-AS1
AL356752.1→
MCF2L→
←AL139384.1
←AL356740.1
AL356740.2→
ATP11A→
←MCF2L-AS1
AL356740.3→

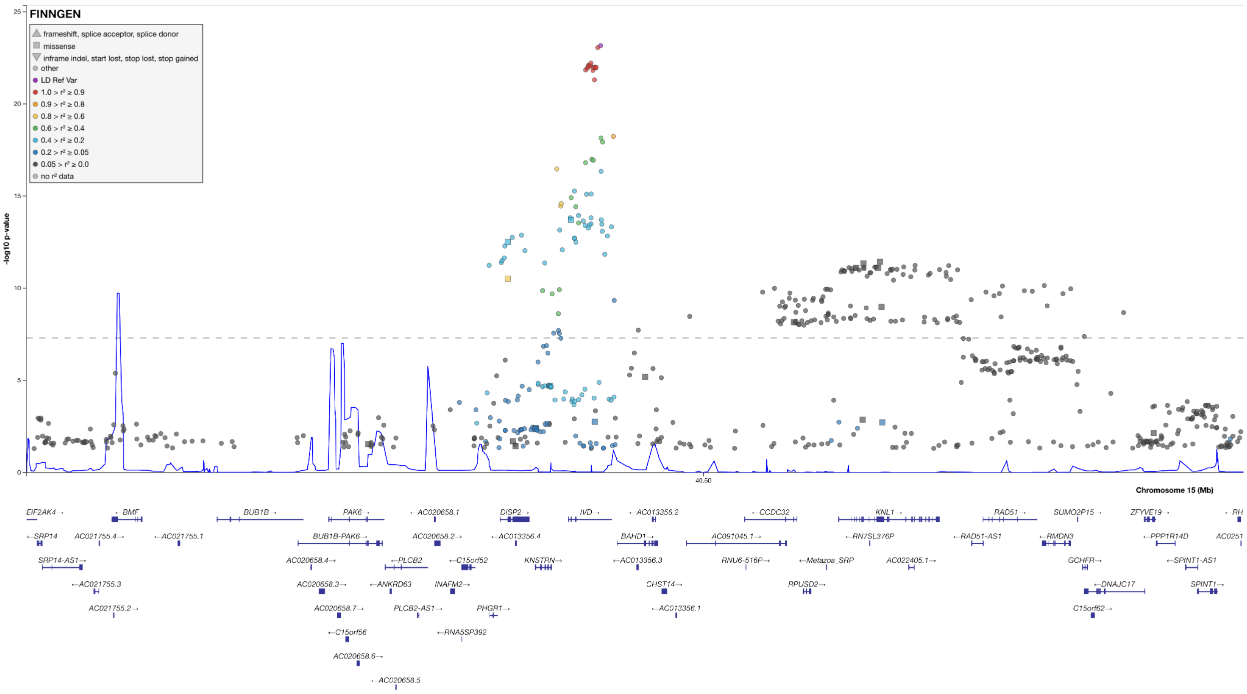
FINNGEN
frameshift, splice acceptor, splice donor
missense
inframe indel, start lost, stop lost, stop gained
other
LD Ref Var
1.0 > r² ≥ 0.9
0.9 > r² ≥ 0.8
0.8 > r² ≥ 0.6
0.6 > r² ≥ 0.4
0.4 > r² ≥ 0.2
0.2 > r² ≥ 0.05
0.05 > r² ≥ 0.0
no r² data
-log10 p-value
25
20
15
10
5
0
40.50
Chromosome 15 (Mb)
EIF2AK4
BMF
BUB1B
PAK6
AC020658.1
DISP2
IVD
AC013356.2
CCDC32
KNL1
RAD51
SUMO2P15
ZFYVE19
SRP14
AC021755.4
AC021755.1
BUB1B-PAK6
AC020658.2
AC013356.4
BAHD1
AC091045.1
RN7SL376P
RAD51-AS1
RMDN3
PPP1R14D
SRP14-AS1
AC020658.4
PLCB2
C15orf52
KNSTRN
AC013356.3
RNU6-516P
Metazoa_SRP
AC022405.1
GCHFR
SPINT1-AS1
AC021755.3
AC020658.3
ANKRD63
INAFM2
CHST14
RPUSD2
DNAJC17
SPINT1
AC021755.2
AC020658.7
PLCB2-AS1
PHGR1
AC013356.1
C15orf62
C15orf56
RNA5SP392
AC020658.6
AC020658.5

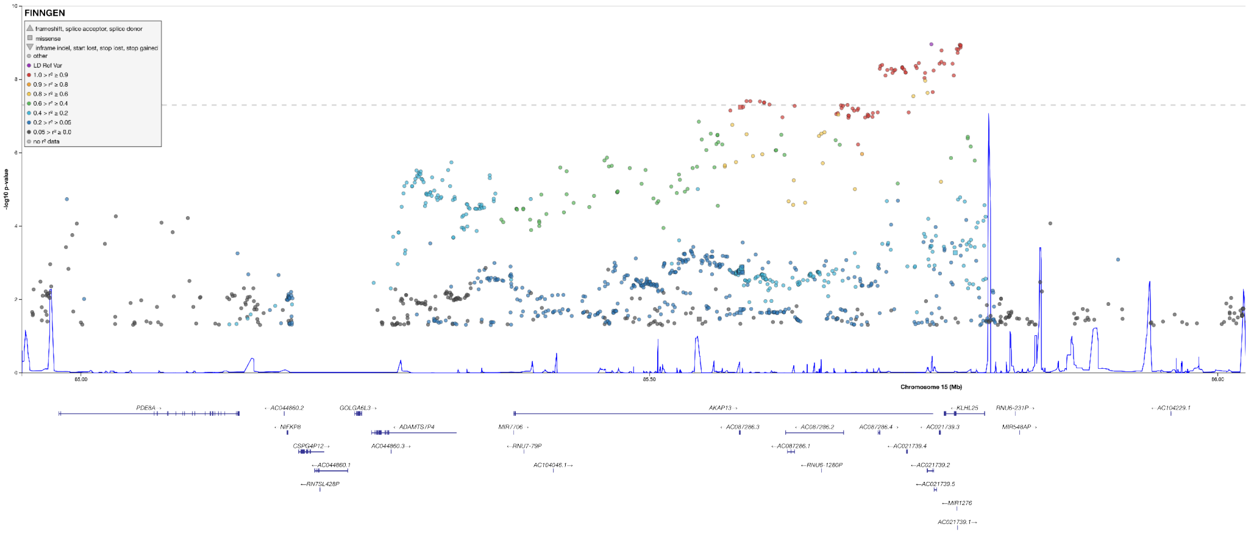
FINNGEN
frameshift, splice acceptor, splice donor
missense
inframe indel, start lost, stop lost, stop gained
other
LD Ref Var
1.0 > r² ≥ 0.9
0.9 > r² ≥ 0.8
0.8 > r² ≥ 0.6
0.6 > r² ≥ 0.4
0.4 > r² ≥ 0.2
0.2 > r² ≥ 0.05
0.05 > r² ≥ 0.0
no r² data
-log10 p-value
10
8
6
4
2
0
85.00
85.50
86.00
Chromosome 15 (Mb)
PDE8A
AC044860.2
GOLGA6L3
AKAP13
KLHL25
RNU6-231P
AC104229.1
NIFKP8
ADAMTSP4
MIR1706
AC087286.3
AC087286.2
AC087286.4
AC021739.3
MIR548AP
CSPG4P12
AC044860.3
RNU7-79P
AC087286.1
AC021739.4
AC044860.1
AC104046.1
RNU6-1280P
AC021739.2
RN7SL428P
AC021739.5
MIR1276
AC021739.1

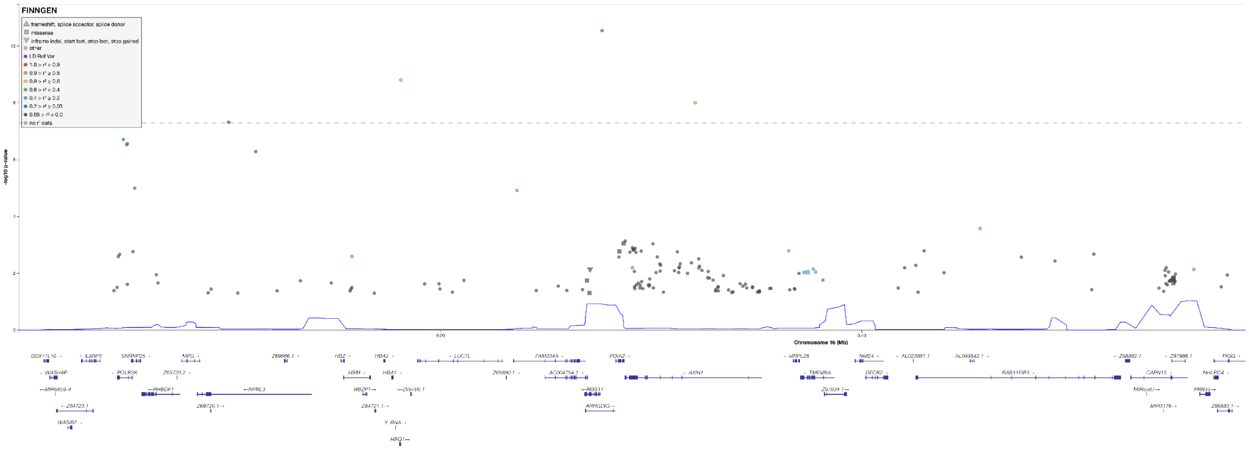
FINNGEN
-log10 p-value
Chromosome 16 (Mb)
AXIN1
RAB11FIP3

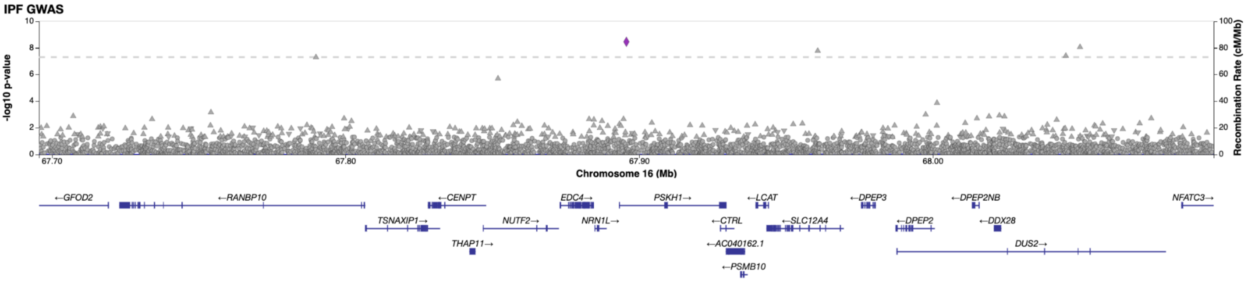
IPF GWAS
-log10 p-value
Recombination Rate (cM/Mb)
Chromosome 16 (Mb)
67.70
67.80
67.90
68.00
←GFOD2
←RANBP10
←CENPT
EDC4→
PSKH1→
←LCAT
←DPEP3
←DPEP2NB
NFATC3→
TSNAXIP1→
NUTF2→
NRN1L→
←CTRL
←SLC12A4
←DPEP2
←DDX28
THAP11→
←AC040162.1
DUS2→
←PSMB10

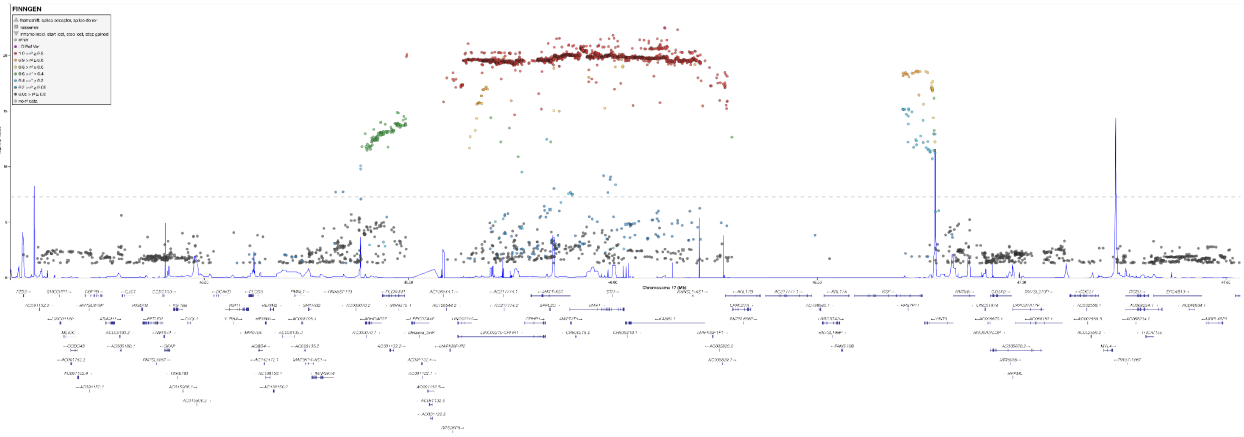
FINNGEN

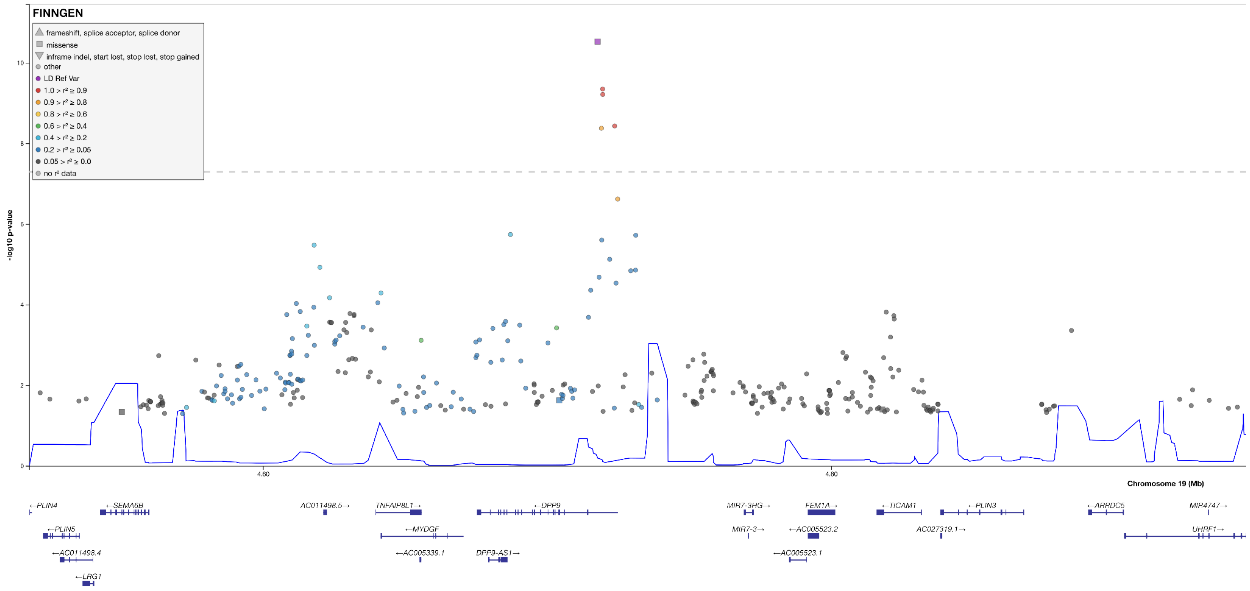
FINNGEN
frameshift, splice acceptor, splice donor
missense
inframe indel, start lost, stop lost, stop gained
other
LD Ref Var
1.0 > r² ≥ 0.9
0.9 > r² ≥ 0.8
0.8 > r² ≥ 0.6
0.6 > r² ≥ 0.4
0.4 > r² ≥ 0.2
0.2 > r² ≥ 0.05
0.05 > r² ≥ 0.0
no r² data
-log10 p-value
Chromosome 19 (Mb)
4.60
4.80
←PLIN4
←SEMA6B
AC011498.5→
TNFAIP8L1→
←DPP9
MIR7-3HG→
FEM1A→
←TICAM1
←PLIN3
←ARRDC5
MIR4747→
←PLIN5
←MYDGF
MIR7-3→
←AC005523.2
AC027319.1→
UHRF1→
←AC011498.4
←AC005339.1
DPP9-AS1→
←AC005523.1
←LRG1

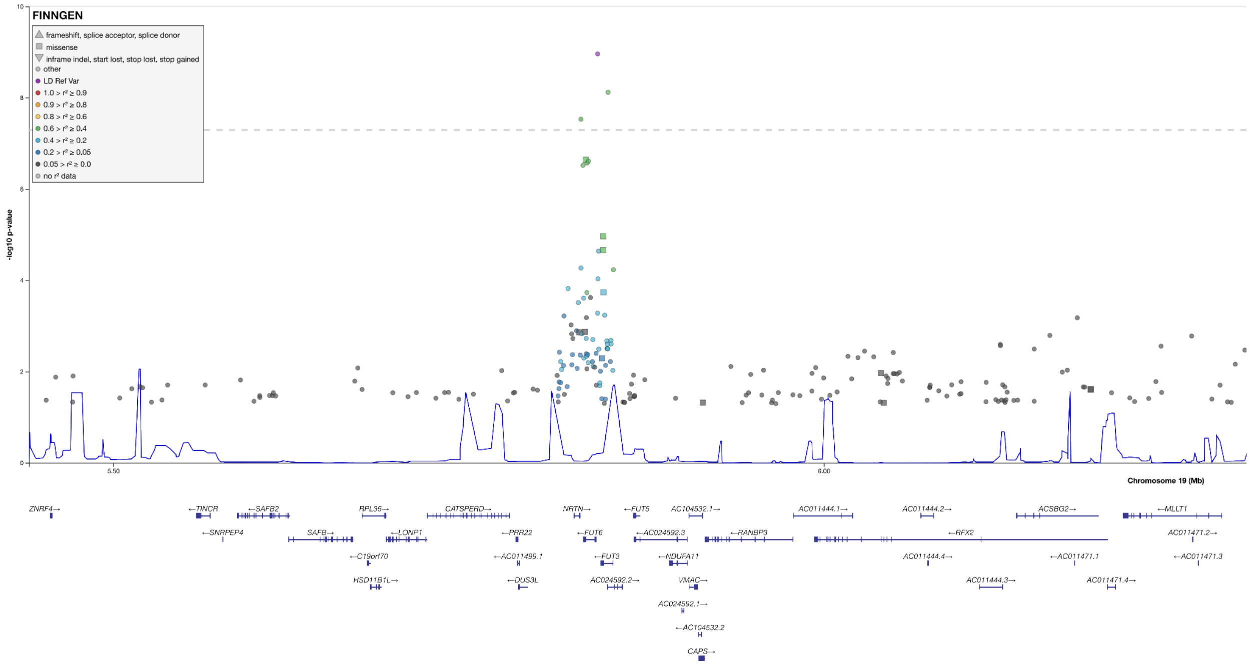
FINNGEN
frameshift, splice acceptor, splice donor
missense
inframe indel, start lost, stop lost, stop gained
other
LD Ref Var
1.0 > r² ≥ 0.9
0.9 > r² ≥ 0.8
0.8 > r² ≥ 0.6
0.6 > r² ≥ 0.4
0.4 > r² ≥ 0.2
0.2 > r² ≥ 0.05
0.05 > r² ≥ 0.0
no r² data
-log10 p-value
Chromosome 19 (Mb)
ZNRF4→
←TINCR
←SAFB2
RPL36→
CATSPERD→
NRTN→
←FUT5
AC104532.1→
AC011444.1→
AC011444.2→
ACSBG2→
←MLLT1
←SNRPEP4
SAFB→
←LONP1
←PRR22
←FUT6
←AC024592.3
←RANBP3
←RFX2
AC011471.2→
←C19orf70
←AC011499.1
←FUT3
←NDUFA11
AC011444.4→
←AC011471.1
←AC011471.3
HSD11B1L→
←DUS3L
AC024592.2→
VMAC→
AC011444.3→
AC011471.4→
AC024592.1→
←AC104532.2
CAPS→

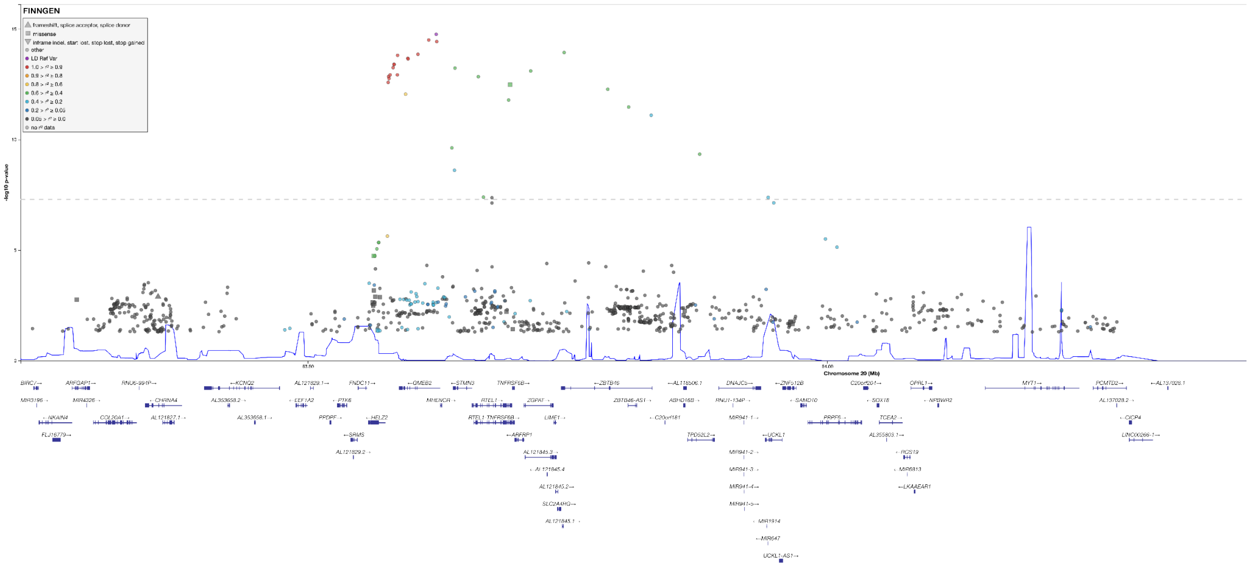
FINNGEN
-log10 p-value
Chromosome 20 (Mb)

**Fig S2.** Quantile-quantile plots for IPF meta-analyses.

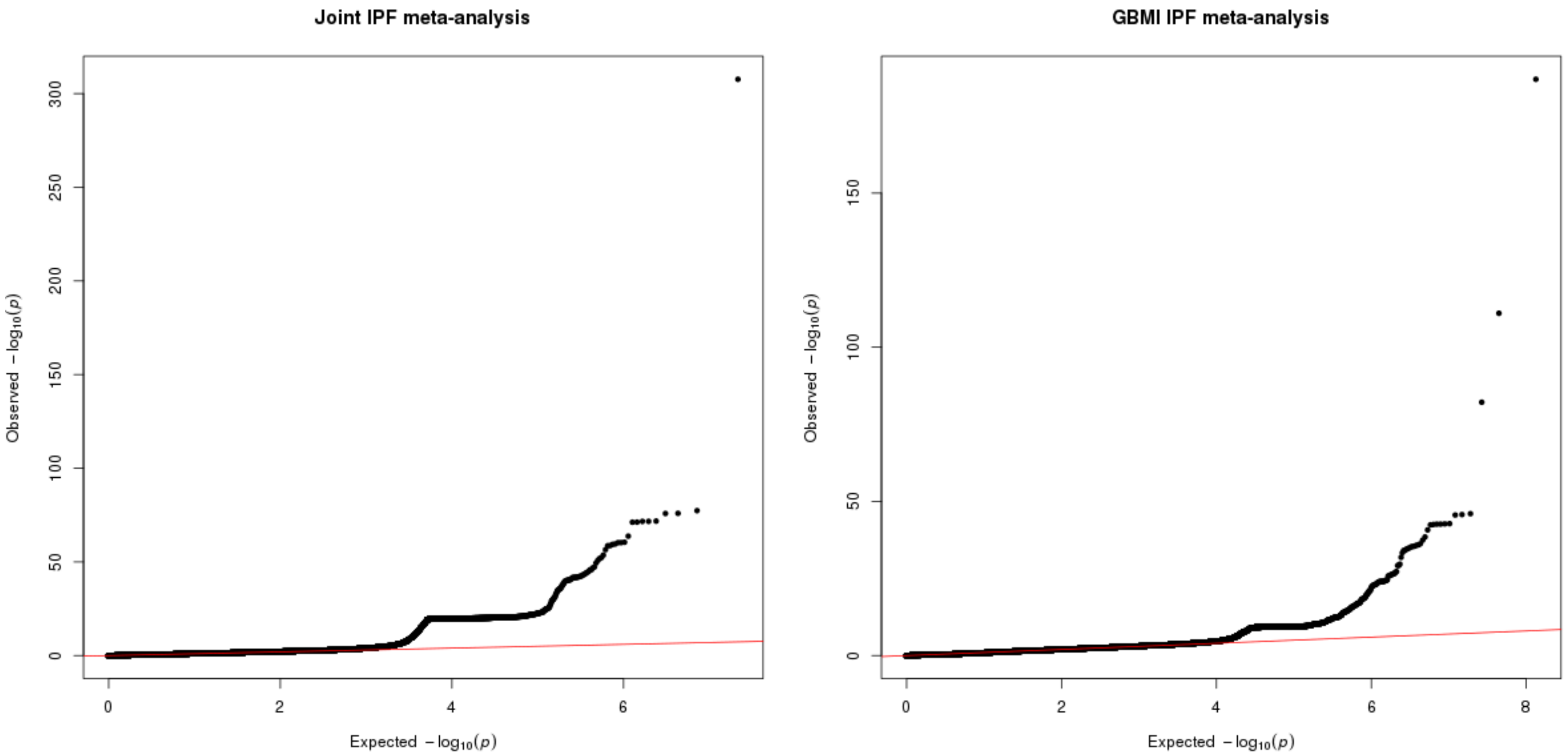

**Fig S3.** Forest plot of effects of *MUC5B* variant rs35705950 by sex for five biobanks with sex-specific results for both males and females and meta-analysis results of these five biobanks.

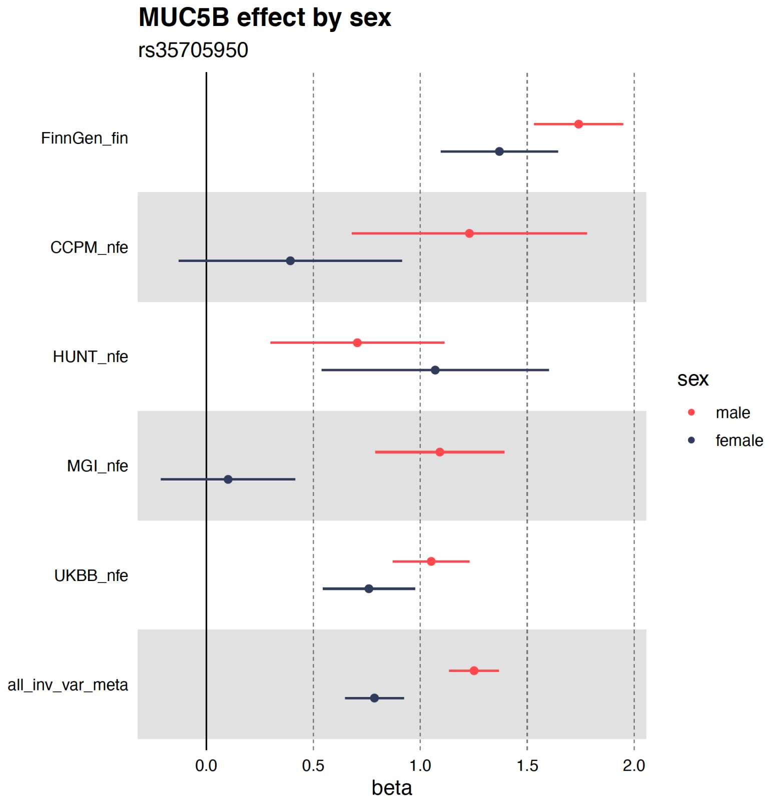

**Fig S4.** Forest plots for all genome-wide significant loci.

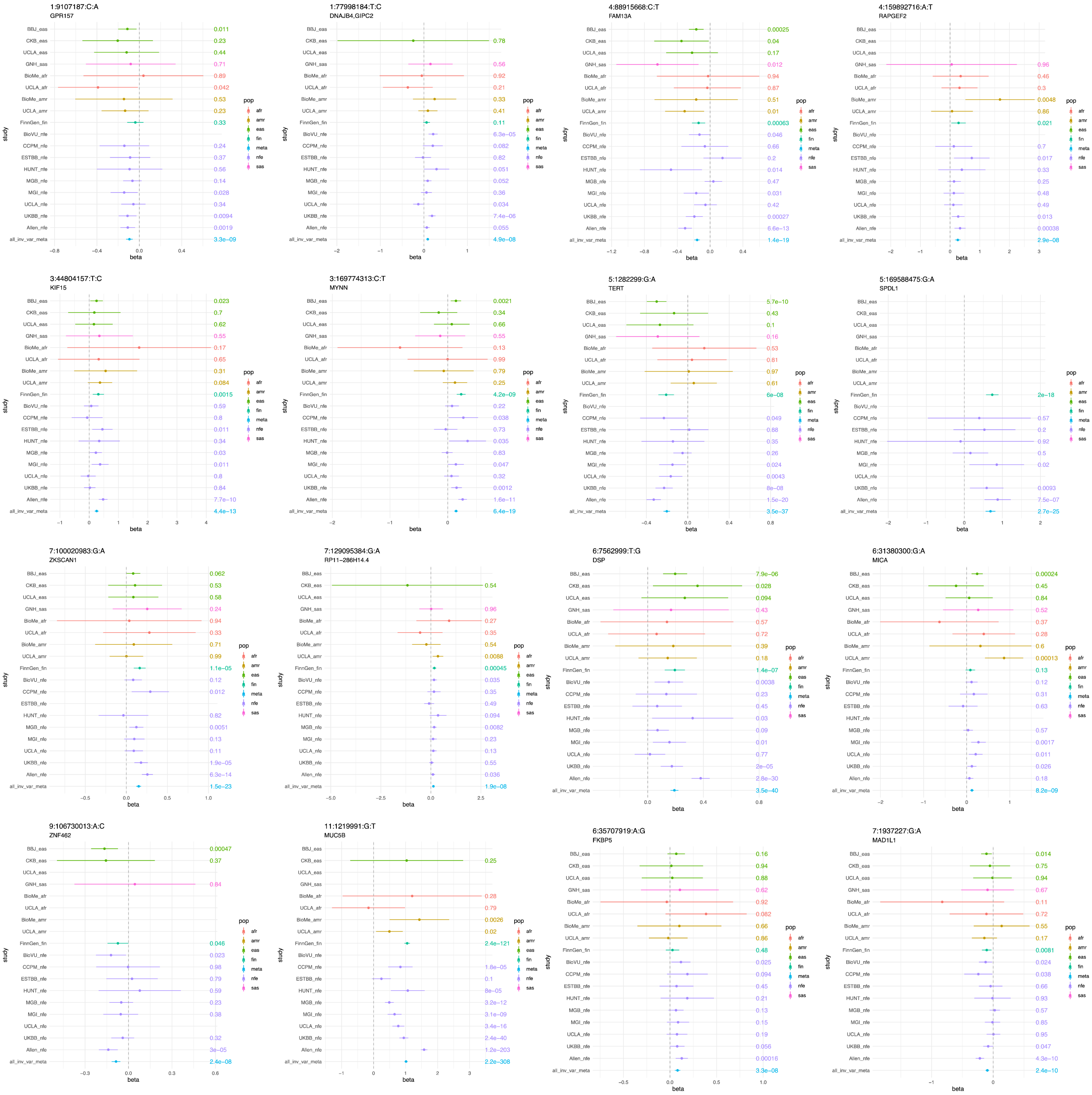

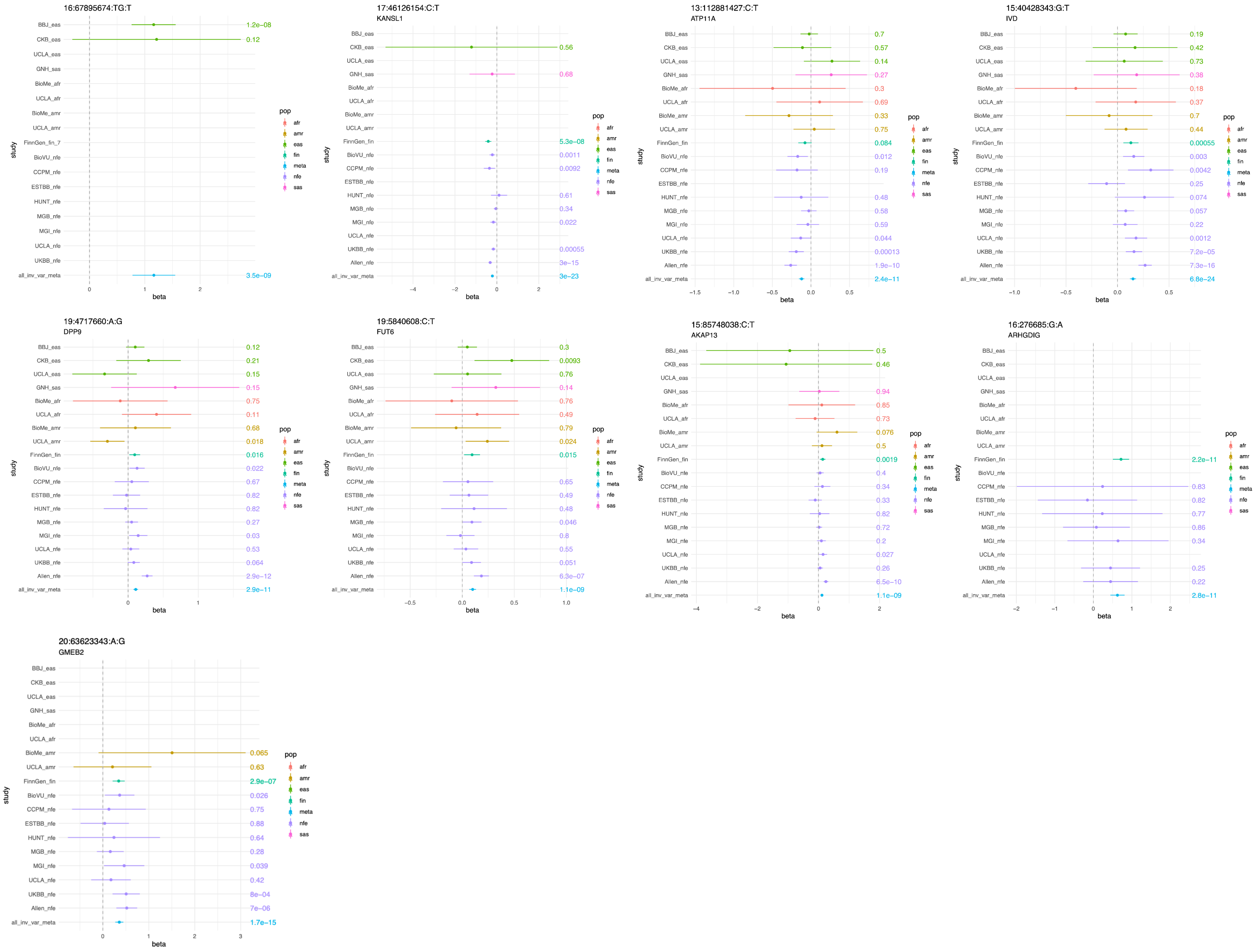
16:67895674:TG:T
17:46126154:C:T
KANSL1
13:112881427:C:T
ATP11A
15:40428343:G:T
IVD
19:4717660:A:G
DPP9
19:5840608:C:T
FUT6
15:85748038:C:T
AKAP13
16:276685:G:A
ARHGDIG
20:63623343:A:G
GMEB2
study
beta
pop
afr
amr
eas
fin
meta
nfe
sas

**Fig S5.** Effect size estimate comparisons GBMI vs. latest IPF meta-analysis (Allen et al.) by biobank.

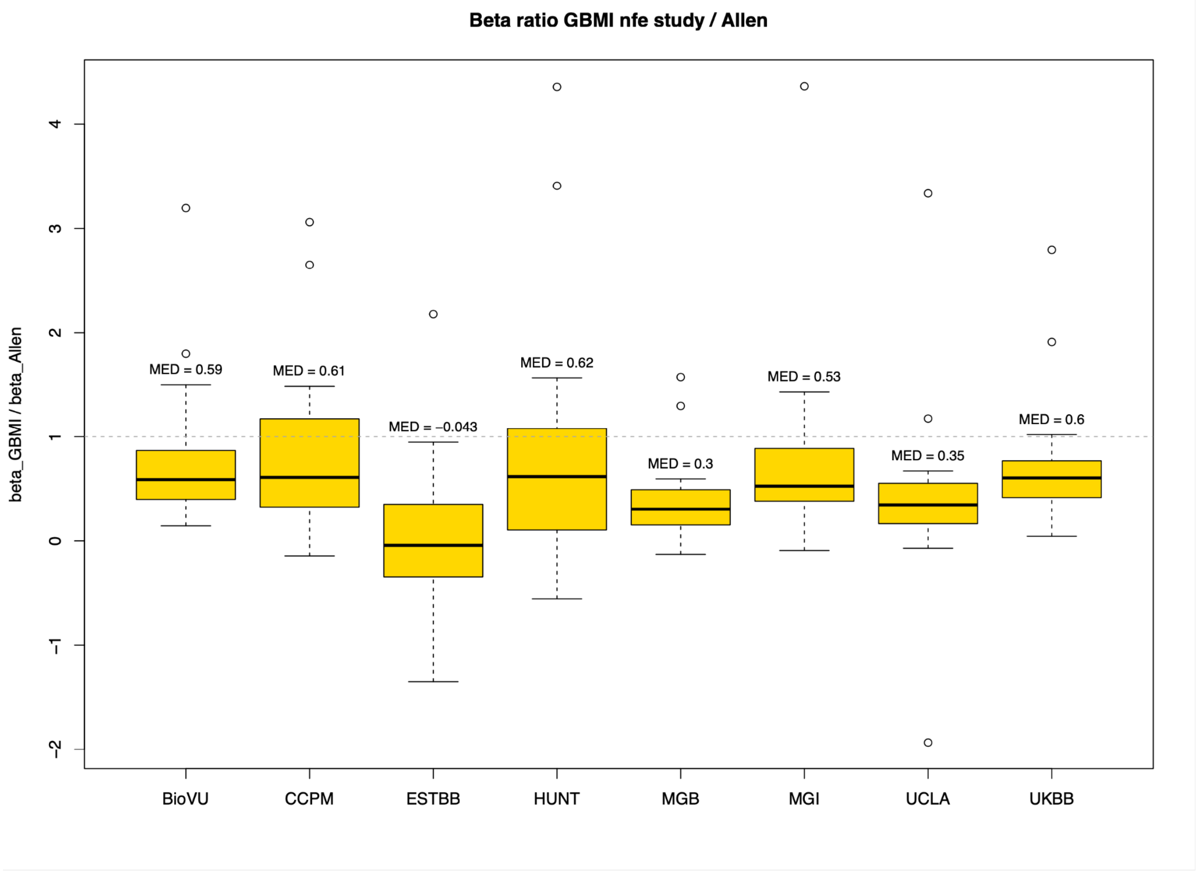

**Fig S6.** Effect size estimate comparison FinnGen subsets vs. latest IPF meta-analysis (Allen et al.). Variants included in the analysis were genome-wide significant in the joint meta-analysis and had the same direction of effects in the respective FinnGen subset and the Allen et al. study.

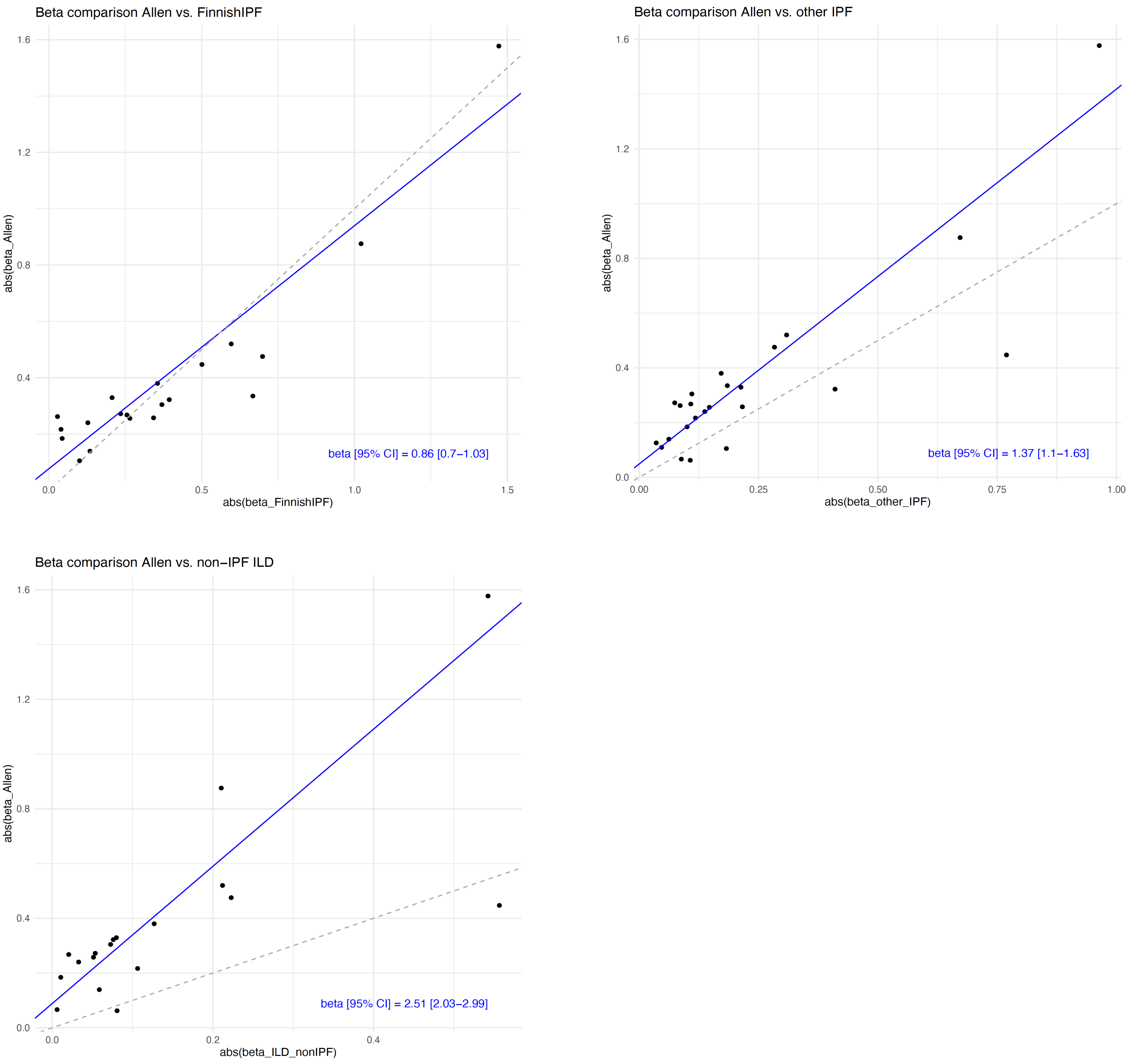

#### Supplementary Tables

**Table S1. Summary of biobanks in GBMI IPF analysis.** *Sex-stratified analyses were conducted in release 5 (as opposed to release 7 here) with 378 and 110524, and 650 and 86462 female and male IPF cases and controls, respectively. **Corresponds to values for entire biobanks, not per ancestry.

| Biobank | Ancestry | IPF patients | Controls | Frac cases (%) | Frac Female (%) | Frac female cases (%) | Mean age (SD) | Sampling strategy |
|---|---|---|---|---|---|---|---|---|
| BBJ | EAS | 1046 | 176974 | 0.59 | 46.3 | 40.7 | 63.1 (9.6) | Hospital/Health center-based |
| BioMe | AFR | 93 | 7218 | 1.27 | 62.6 | 76.3 | 50.5 (17.6)** | Hospital/Health center-based |
| BioMe | AMR | 115 | 10281 | 1.11 | 63.1 | 64.3 | 50.5 (17.6)** | Hospital/Health center-based |
| BioVU | NFE | 745 | 49259 | 1.49 | 59.6 | 48.6 | NA | Hospital/Health center-based |
| CCPM | NFE | 163 | 21694 | 0.75 | 64.0 | 49.1 | 50.3 (17.5) | Hospital/Health center-based |
| CKB | EAS | 75 | 75224 | 0.10 | NA | NA | 53.7 (11.0) | Population-based |
| ESTBB | NFE | 250 | 133968 | 0.19 | NA | NA | NA | Population-based |
| FinnGen* | FIN | 1514 | 306063 | 0.49 | 56.3 | 35.5 | 59.1 (17.9) | Mixed |
| GNH | SAS | 51 | 21897 | 0.23 | NA | NA | 41.2 (13.8) | Population-based |
| HUNT | NFE | 235 | 64845 | 0.36 | 53.4 | 40.4 | 69.9 (18.4) | Population-based |
| MGB | NFE | 1305 | 16628 | 7.28 | NA | NA | 60 (16.5) | Hospital/Health center-based |
| MGI | NFE | 566 | 43611 | 1.28 | 54.1 | 44.7 | 56.9 (16.4) | Hospital/Health center-based |
| UCLA | AFR | 76 | 1150 | 6.20 | NA | NA | 56.4 (17.3)** | Hospital/Health center-based |
| UCLA | AMR | 204 | 4171 | 4.66 | NA | NA | 56.4 (17.3)** | Hospital/Health center-based |
| UCLA | EAS | 89 | 2211 | 3.87 | NA | NA | 56.4 (17.3)** | Hospital/Health center-based |
| UCLA | NFE | 700 | 15941 | 4.21 | NA | NA | 56.4 (17.3)** | Hospital/Health center-based |
| UKBB | NFE | 1265 | 404684 | 0.31 | 55.0 | 38.8 | NA | Population-based |
| **13** | **6** | **8492** | **1355819** | **0.62** | | | | |

**Table S2. Results for all genome-wide significant loci, including heterogeneity statistics per locus: Cochran's Q p-value for heterogeneity (Q_pval) and heterogeneity index (I²).**

| SNPID | nearest gene | most severe consequence | OR[95% CI] | meta_pval | meta_N_studies | I² | Q_pval | Q_pval_FDR_adjusted |
|---|---|---|---|---|---|---|---|---|
| 1:9107187:C:A | *GPR157* | intron | 0.91 [0.88–0.94] | 3.28E-09 | 17 | 0 | 0.9657 | 0.9657 |
| 1:77998184:T:C | *DNAJB4* | intron | 1.09 [1.06–1.13] | 4.91E-08 | 16 | 0.534 | 0.0061 | 0.0174 |
| 3:44804157:T:C | *KIF15* | intron | 1.29 [1.2–1.38] | 4.37E-13 | 18 | 0.358 | 0.0663 | 0.1184 |
| 3:169774313:C:T | *MYNN* | synonymous | 1.16 [1.12–1.19] | 6.43E-19 | 18 | 0.590 | 8.09E-04 | 0.0029 |
| 4:88915668:C:T | *FAM13A* | intron | 0.85 [0.82–0.88] | 1.36E-19 | 18 | 0.624 | 2.30E-04 | 0.0014 |
| 4:159892716:A:T | *RAPGEF2* | intergenic | 1.29 [1.18–1.42] | 2.94E-08 | 14 | 0 | 0.5195 | 0.6493 |
| 5:1282299:G:A | *TERT* | intron | 0.82 [0.79–0.84] | 3.56E-37 | 17 | 0.651 | 1.03E-04 | 8.55E-04 |
| 5:169588475:G:A | *SPDL1* | missense | 1.99 [1.75–2.27] | 2.68E-25 | 8 | 0.091 | 0.3597 | 0.4996 |
| 6:7562999:T:G | *DSP* | intron | 1.21 [1.18–1.25] | 3.53E-40 | 18 | 0.699 | 3.91E-06 | 4.89E-05 |
| 6:31380300:G:A | *MICA* | upstream gene | 1.13 [1.08–1.18] | 8.17E-09 | 17 | 0.418 | 0.0364 | 0.0758 |
| 6:35707919:A:G | *FKBP5* | intron | 1.08 [1.05–1.12] | 3.33E-08 | 18 | 0 | 0.9286 | 0.9657 |
| 7:1937227:G:A | *MAD1L1* | intron | 0.91 [0.88–0.94] | 2.35E-10 | 18 | 0.451 | 0.0201 | 0.0457 |
| 7:100020983:G:A | *ZKSCAN1* | intron | 1.16 [1.13–1.2] | 1.52E-23 | 17 | 0.273 | 0.1433 | 0.2388 |
| 7:129095384:G:A | *RP11-286H 14.4* | non coding transcript exon | 1.13 [1.09–1.19] | 1.94E-08 | 16 | 0 | 0.4838 | 0.6366 |
| 9:106730013:A:C | *ZNF462* | intron | 0.92 [0.89–0.95] | 2.41E-08 | 12 | 0.089 | 0.3584 | 0.4996 |
| 11:1219991:G:T | *MUC5B* | upstream gene variant | 2.76 [2.62–2.9] | 0 | 14 | 0.944 | 6.50E-42 | 1.62E-40 |
| 13:112881427:C:T | *ATP11A* | non coding transcript exon | 0.89 [0.86–0.92] | 2.35E-11 | 17 | 0.515 | 0.0073 | 0.0184 |
| 15:40428343:G:T | *IVD* | intron | 1.16 [1.13–1.2] | 6.82E-24 | 18 | 0.514 | 0.0062 | 0.0174 |

| | | | | | | | | |
|---|---|---|---|---|---|---|---|---|
| 15:85748038:C:T | *AKAP13* | 3 prime UTR | 1.12 [1.08–1.16] | 1.10E-09 | 17 | 0.384 | 0.0545 | 0.1049 |
| 16:276685:G:A | *ARHGDIG* | intron | 1.87 [1.55–2.25] | 2.85E-11 | 8 | 0 | 0.7230 | 0.8216 |
| 16:67895674:TG:T | *PSKH1* | intron | 3.2 [2.17–4.71] | 3.52E-09 | 2 | 0 | 0.9470 | 0.9657 |
| 17:46126154:C:T | *KANSL1* | intron | 0.8 [0.77–0.84] | 3.00E-23 | 10 | 0.696 | 5.21E-04 | 0.0026 |
| 19:4717660:A:G | *DPP9* | missense | 1.12 [1.08–1.15] | 2.94E-11 | 18 | 0.597 | 6.34E-04 | 0.0026 |
| 19:5840608:C:T | *FUT6* | upstream gene | 1.11 [1.07–1.14] | 1.08E-09 | 17 | 0.150 | 0.2775 | 0.4336 |
| 20:63623343:A:G | *GMEB2* | intron | 1.43 [1.31–1.56] | 1.69E-15 | 12 | 0 | 0.5788 | 0.6891 |

**Table S3. Replication of previously reported IPF associated variants in joint IPF meta-analysis.** Loci which are not significant after conditioning on MUC5B are not included. *rs7144383 was the only variant with opposite directions of effects in this study compared to previous findings ** GBMI meta-analysis results (variant missing from joint meta-analysis).

| rsid | variant | locus | Effect | P-value | variant analyzed (LD) | Study |
|---|---|---|---|---|---|---|
| rs78238620 | chr3_44860894_T_A | KIF15 | 0.24 | 9.59E-13 | actual | Allen et al. |
| rs12696304 | chr3_16976348_C_G | ACTRT3 | 0.14 | 6.4E-19 | rs10936599 ($r^2$ = 0.86) | Fingerlin et al. |
| rs2013701 | chr4_88963935_G_T | FAM13A | -0.12 | 9.69E-17 | actual | Fingerlin et al. |
| rs7725218 | chr5_1282299_G_A | TERT | -0.20 | 3.55E-37 | actual | Mushiroda et al. |
| rs116483731 | chr5_169588475_G_A | SPDL1 | 0.69 | 2.68E-25 | actual | Dhindsa et al. |
| rs2076295 | chr6_7562999_T_G | DSP | 0.19 | 3.53E-40 | actual | Fingerlin et al. |
| rs7887 | chr6_31896770_G_T | EHMT2 | -0.03 | 0.0254585 | actual | Fingerlin et al. 2016 |
| rs2897075 | chr7_100032719_C_T | ZKSCAN1 | 0.15 | 5.98E-23 | actual | Fingerlin et al. |
| rs12699415 | chr7_1869843_A_G | MAD1L1 | -0.08 | 5.03E-07 | actual | Allen et al. |
| rs28513081 | chr8_119921886_A_G | DEPTOR | -0.07 | 1.20E-05 | actual | Allen et al. |
| rs6477542 | chr9_106745151_C_T | ZNF462 | 0.09 | 9.87E-07 | actual | Ishigaki et al. |
| rs537322302 | chr10_93271016_C_G | HECTD2 | NA | NA | NA | Allen et al. |
| rs11191865 | chr10_103913084_G_A | OBFC1 | 0.06 | 1.26E-04 | actual | Fingerlin et al. |
| rs967235139 | chr10_122092604_T_C | TACC2 | 1.1141** | 6.85E-05* | actual | Koskela et al. |
| rs35705950 | chr11_1219991_G_T | MUC5B | 1.01 | < 1E-300 | actual | Seibold et al. |
| rs9577395 | chr13_112880670_C_G | ATP11A | -0.11 | 3.39E-10 | actual | Fingerlin et al. |
| rs7144383* | chr14_47571172_G_A | MDGA2 | -0.02 | 0.39936458 | actual | Noth et al. |
| rs59424629 | chr15_40428343_G_T | IVD | 0.15 | 6.82E-24 | actual | Fingerlin et al. |
| rs62023891 | chr15_85553985_G_A | AKAP13 | 0.09 | 3.01E-07 | actual | Allen et al. 2017 |
| rs185488877 | chr16_344551_C_T | AXIN1 | 0.61** | 1.65E-10 | actual | Koskela et al. |
| rs2077551 | chr17_46137522_T_C | MAPT | -0.21 | 2.57E-14 | rs113120855 ($r^2$ = 0.85) | Noth et al. / Fingerlin et al. |
| rs12610495 | chr19_4717660_A_G | DPP9 | 0.11 | 2.95E-11 | actual | Fingerlin et al. |
| rs41308092 | chr20_63693038_G_A | RTEL1 | 0.35 | 1.60E-12 | actual | Allen et al. |

**Table S4. European meta-analysis results for novel loci.** European meta-analysis was limited to FIN and NFE ancestries.

| SNPID | nearest gene | most severe consequence | OR[95% CI] | meta_pval | meta_N _studies | meta_Q_pval | meta_I2 |
|---|---|---|---|---|---|---|---|
| 1:9107187:C:A | *GPR157* | intron | 0.92[0.89-0.95] | 9.68E-07 | 9 | 0.87795 | 0 |
| 1:77998184:T:C | *DNAJB4* | intron | 1.09[1.06-1.13] | 6.39E-08 | 10 | 0.00063 | 0.69043177 |
| 4:159892716:A:T | *RAPGEF2* | intergenic | 1.28[1.17-1.41] | 1.76E-07 | 9 | 0.65049 | 0 |
| 6:35707919:A:G | *FKBP5* | intron | 1.09[1.05-1.12] | 1.03E-07 | 10 | 0.73352 | 0 |
| 7:129095384:G:A | *RP11-286H14.4* | non coding transcript exon | 1.13[1.08-1.18] | 9.45E-08 | 10 | 0.54158 | 0 |
| 16:67895674:TG:T | *PSKH1* | intron | NA | NA | 0 | NA | NA |
| 19:5840608:C:T | *FUT6* | upstream gene | 1.11[1.07-1.15] | 3.34E-08 | 9 | 0.29386 | 0.16708471 |

**Table S5. Replication of novel loci in two independent European cohorts.**

| | | | | | | | | | **UUS** | | | | **Genentech** | | | **UUS Genentech meta-analysis** | |
|---|---|---|---|---|---|---|---|---|---|---|---|---|---|---|---|---|---|
| **chr** | **pos** | **ref** | **alt** | **rsid** | **nearest gene** | **AF (%)** | **beta** | **pval** | **IMPUTATION R2** | **AF (%)** | **beta** | **pval** | **AF (%)** | **beta** | **pval** | **beta** | **pval** |
| 1 | 77998184 | T | C | rs4130548 | *DNAJB4, GIPC2* | 33.28 | 0.088 | 4.91E-08 | 0.99 | 38.04 | 0.100 | 0.082 | 35.13 | -0.030 | 0.653 | 0.046 | 0.297 |
| 1 | 9107187 | C | A | rs7549256 | *GPR157* | 64.15 | -0.094 | 3.29E-09 | 0.98 | 66.74 | -0.233 | 5.26E-05 | 66.83 | -0.180 | 0.011 | -0.212 | 2.08E-06 |
| 4 | 159892716 | A | T | rs76537958 | *RAPGEF2* | 2.923 | 0.258 | 2.94E-08 | 0.99 | 3.44 | -0.178 | 0.277 | 3.57 | -0.159 | 0.384 | -0.169 | 0.164 |
| 6 | 35707919 | A | G | rs9380529 | *FKBP5* | 51.73 | 0.081 | 3.33E-08 | 1.00 | 48.99 | 0.131 | 0.020 | 48.46 | 0.099 | 0.134 | 0.117 | 0.0062 |
| 7 | 129095384 | G | A | rs34288126 | *RP11-286H14.4* | 12.7 | 0.127 | 1.50E-08 | 0.99 | 13.12 | 0.111 | 0.169 | 13.52 | 0.014 | 0.881 | 0.071 | 0.252 |
| 16 | 67895674 | TG | T | rs539683219 | *PSKH1* | 1.709 | 1.163 | 3.52E-09 | NA | NA | NA | NA | NA | NA | NA | NA | NA |
| 19 | 5840608 | C | T | rs708686 | *FUT6* | 31.48 | 0.100 | 1.08E-09 | 0.99 | 26.82 | 0.150 | 0.014 | 30.81 | 0.079 | 0.268 | 0.120 | 0.0097 |

**Table S6. COVID-19 hospitalized vs population genome-wide significant lead variants reaching FDR-adjusted p-value < 0.05 in joint IPF meta-analysis (6/17 35%)**

| SNP | rsid | nearest gene | most severe consequnce | beta IPF | beta covid | p IPF | p covid |
|---|---|---|---|---|---|---|---|
| 6:31153455:T:C | rs111837807 | *CCHCR1* | intron | -0.0786433 | 0.12021 | 0.00224954 | 2.33E-11 |
| 11:1219991:G:T | rs35705950 | *MUC5B* | upstream_gene | 1.0137895 | -0.11154 | <1E-300 | 6.47E-09 |
| 16:89196249:G:A | rs117169628 | *SLC22A31* | missense | 0.07015378 | 0.088157 | 0.00147992 | 2.55E-08 |
| 17:45707983:T:C | rs61667602 | *CRHR1* | intron | -0.1866383 | -0.093206 | 2.54E-20 | 3.77E-12 |
| 17:49863303:C:T | rs77534576 | *TAC4* | intergenic | 0.10690078 | 0.20867 | 0.00621308 | 4.29E-11 |
| 19:4719431:G:A | rs2109069 | *DPP9* | intron | 0.10143175 | 0.11214 | 4.40E-10 | 2.05E-22 |

**Table S7. Fraction of females and MUC5B (rs35705950) estimates in biobanks.**

| Biobank | N_female_cases | N_male_cases | N_female_controls | N_male_controls | Frac_female_cases | Frac_female | *MUC5B* OR [95% CI] | *MUC5B* pval |
|---|---|---|---|---|---|---|---|---|
| MGI_nfe | 253 | 313 | 23660 | 19951 | 0.447 | 0.541 | 1.93[1.55–2.39] | 3.11E-09 |
| CCPM_nfe | 80 | 83 | 13899 | 7795 | 0.491 | 0.64 | 2.31[1.58–3.39] | 1.77E-05 |
| UKBB_nfe | 491 | 782 | 222681 | 185194 | 0.386 | 0.545 | 2.57[2.23–2.95] | 2.43E-40 |
| FinnGen_fin | 378 | 650 | 110524 | 86462 | 0.368 | 0.56 | 2.86[2.62–3.12] | 0 |
| HUNT_nfe | 95 | 140 | 34644 | 30201 | 0.404 | 0.534 | 2.91[1.71–4.94] | 7.98E-05 |
| BioMe_afr | 71 | 0 | 4506 | 0 | 1 | 1 | 3.33[0.38–29.26] | 2.77E-01 |
| BioMe_amr | 74 | 0 | 6484 | 0 | 1 | 1 | 4.18[1.65–10.59] | 2.58E-03 |
| BBJ_eas | 426 | 620 | 82081 | 94893 | 0.407 | 0.463 | NA | NA |
| BioVU_nfe | 362 | 383 | 29456 | 19803 | 0.486 | 0.596 | NA | NA |

**Table S8. Proportion of lung transplants or IPF-specific deaths among IPF cases in FinnGen by *MUC5B* rs35705950 risk allele carrier status.** Fisher's exact test estimates from a dominant model where risk allele heterozygotes and homozygotes are grouped together.

| | **All** | | **Women** | | **Men** | |
|---|---|---|---|---|---|---|
| | **MUC5B** | **no MUC5B** | **MUC5B** | **no MUC5B** | **MUC5B** | **no MUC5B** |
| **N in FinnGen** | 59748 | 249406 | 33530 | 140216 | 26218 | 109190 |
| % | 19.3 % | 80.7 % | 19.3 % | 80.7 % | 19.4 % | 80.6 % |
| **IPF diagnosis** | | | | | | |
| N | 685 | 828 | 229 | 310 | 456 | 518 |
| % | 1.15 % | 0.33 % | 0.68 % | 0.22 % | 1.74 % | 0.47 % |
| p | p<2.2e-16 | | p<2.2e-16 | | p<2.2e-16 | |
| OR | 3.48 (3.41 - 3.86) | | 3.10 (2.60 - 3.70) | | 3.71 (3.26 - 4.22) | |
| **Subsequent lung transplant or IPF death** | | | | | | |
| N | 162 | 201 | 41 | 59 | 121 | 142 |
| **% of all in FinnGen** | 0.27 % | 0.08 % | 0.12 % | 0.04 % | 0.46 % | 0.13 % |
| p | p<2.2e-16 | | p=6.3e-7 | | p<2.2e-16 | |
| OR | 3.37 (2.72 - 4.17) | | 2.91 (1.90 - 4.41) | | 3.56 (2.77 - 4.57) | |
| **% of IPF diagnosed** | 23.65 % | 24.28 % | 17.90 % | 19.03 % | 26.54 % | 27.41 % |
| p | p=0.81 | | p=0.82 | | p=0.77 | |
| OR | 0.966 (0.756 - 1.23) | | 0.928 (0.580 - 1.47) | | 0.956 (0.713 - 1.28) | |

**Table S9. Additional genome-wide significant loci identified by sex-stratified meta-analyses**

| sex | CHR | POS (hg38) | REF | ALT (Effect) | rsid | Function | Gene | AF (ALT) | beta (ALT) | se | p-value | Q_pval | direction ALT | N BB | Q_pval btw sexes |
|---|---|---|---|---|---|---|---|---|---|---|---|---|---|---|---|
| Male | 19 | 45641397 | A | AGACCT | rs71338787 | intron | *EML2* | 0.178 | 0.247 | 0.0428 | 8.31E-09 | 0.98 | +??+??+ | 3 | 1.53E-06 |
| Male | 23 | 154784234 | C | T | rs5945238 | intergenic | NONE | 0.072 | 0.207 | 0.0370 | 2.39E-08 | 0.40 | ++?++++ | 6 | 7.50E-05 |

**Table S10. IPF PheCode definition.**

| Trait | PheCode | Participants with these ICD codes are excluded from control population | ICD codes used to define cases |
|---|---|---|---|
| Idiopathic pulmonary fibrosis | 502 | 495,495.0,495.1,495.2,495.3,495.4,495.5,495.6,495.7,495.8,495.9,500,500.0,501,501.0,502,503,504,505,506,506.0,506.1,506.2,506.3,506.4,506.9,507,507.0,507.1,507.8,508,508.0,508.1,508.2,508.8,508.9,510,510.0,510.9,511,511.0,511.1,511.8,511.89,511.9,512,512.0,512.1,512.2,512.8,512.81,512.82,512.83,512.84,512.89,514,514.0,516,516.0,516.1,516.2,516.3,516.30,516.31,516.32,516.33,516.34,516.35,516.36,516.37,516.4,516.5,516.61,516.62,516.63,516.64,516.69,516.8,516.9,517.3,518.0,518.1,518.2,518.3,518.4,518.5,518.51,518.52,518.53,518.81,518.82,518.83,518.84,799.1,V46.1,V46.11,V46.12,V46.13,V46.14,V46.2,B22.1,J18.2,J60,J61,J62,J62.0,J62.8,J63,J63.0,J63.1,J63.2,J63.3,J63.4,J63.5,J63.8,J64,J65,J66,J66.0,J66.1,J66.2,J66.8,J67,J67.0,J67.1,J67.2,J67.3,J67.4,J67.5,J67.6,J67.7,J67.8,J67.9,J68,J68.0,J68.1,J68.2,J68.3,J68.4,J68.8,J68.9,J69.0,J69.1,J69.8,J70,J70.0,J70.1,J70.2,J70.3,J70.4,J70.8,J70.9,J80,J81,J82,J84.0,J84.9,J86,J86.0,J86.9,J90,J91,J92,J92.0,J92.9,J93,J93.0,J93.1,J93.8,J93.9,J94,J94.0,J94.1,J94.2,J94.8,J94.9,J95.1,J95.2,J95.3,J96.0,J96.1,J96.9,J98.1,J98.2,J98.3,R09.1,R09.2,Z99.1,Z99.1,Z99.11,Z99.12,Z99.81,J95.850,J86,J93.12,J86.0,J93.1,J93.81,J86.9,J95.81,J93.11,J93.82,J93.8,J95.812,J95.811,J93.83,J93.0,J93,J93.9,J69.0,J81.1,J18.2,J95.3,J95.822,J95.2,J95.821,J95.82,J95.1,J81.0,J82,D57.811,D57.211,D57.411,D57.01,J98.2,J98.1,J98.11,J98.3,J98.19,R09.1,J91.8,J92,J94.8,J94,J92.0,J94.2,J94.9,J94.0,J90,J94.1,J92.9,J96.9,R06.03,J80,J70.0,J68.3,J70.4,J69.1,J68.8,J70.2,J68.0,J70.5,J68.9,J68.4,J69.8,J70.3,J70.8,J68,J70.1,J68.1,J70,J68.2,J70.9,J84.2,J84.116,J84.11,J84.117,J84.112,J84.114,J84.113,J84.111,J84.115,J96.12,J96.00,J96.20,J96.90,J96.01,J96.10,J96.22,J96.2,J96.02,J96.1,J96.91,J96.92,J96.11,J96.21,J96.0,R09.2,J66.2,J66.0,J67.9,J66.1,J67.8,J66,J67.6,J67,J67.5,J67.7,J67.1,J67.4,J67.3,J67.2,J67.0,J66.8,J63.6,J64,J61,J63.4,J60,J62.8,J63,J63.0,J63.1,J62,J63.5,J62.0,J63.3,J65,J63.2,J84.8,J84.81,J84.09,J84.841,J84.02,J84.82,J84.83,J84.843,J84.0,J84.848,J84.01,J84.03,J84.9,J84.842 | 515,515.0,J84.1,J84.8,J84.89,J84.17,J84.10 |

**Table S11. Phenotype definitions used by biobanks**

| Biobank | Phenotype definition of Idiopathic pulmonary fibrosis (IPF) |
|---|---|
| **Biobank Japan (BBJ)** | Physician's diagnosis OR past medical history (including self-reported)<br>(Possibly include other interstitial lung disease patients.) |
| **BioMe** | phecode |
| **BioVU** | phecode |
| **China Kadoorie Biobank (CKB)** | Cases: J84.1, J84.8; Controls: Exclude B22.1, J18.2, J60-J70, J80-J84, J86, J90-J94,J95.1-J95.3, J96.0, J96.1, J96.9, J98.1-J98.3, R09.1, R09.2, Z99.1 |
| **Colorado Center for Personalized Medicine** | ICD case-control defn from phenotype endpoints |
| **Genes & Health** | ICD10:J841,J849,SNOMED:51615001,90117007,162974009,196125002 |
| **Estonian Biobank** | ICD case-control defn from phenotype endpoints |
| **FinnGen** | J841 |
| **Trøndelag Health Study (HUNT)** | phecode |
| **Michigan Genomics Initiative (MGI)** | phecode |
| **Mass General Brigham** | Cases: ICD-10 J84.1,J84.8; Controls: non-cases excluding anyone with J18.2,J60,J61,J62,J63,J64,J65,J66,J67,J68,J69.0,J69.1,J69.8,J70,J80,J81,J82,J84.0,J84.11,J84.2,J84.8,J84.9,J86,J90,J91,J92,J93,J94,J95.1,J95.2,J95.3,J95.81,J95.82, J95.850,J96.0,J96.1,J96.2,J96.9,J98.1,J98.2,J98.3,D57.211,D57.811,D57.411,D57.01,R09.1,R09.2,Z99.1, Z99.81 |
| **UCLA Precision Health Biobank** | phecode |
| **UK Biobank** | phecode |

**Table S12. Colocalization analysis results in FinnGen.** CLPP = causal posterior probability, CLPA = causal posterior agreement.

| pheno1 | pheno2 | CLPP | CLPA | locus_id1 | locus_id2 |
|---|---|---|---|---|---|
| IPF | ILD | 1 | 1 | 11_1219991_G_T | 11_1219991_G_T |
| IPF | ILD_ENDPOINTS | 1 | 1 | 11_1219991_G_T | 11_1219991_G_T |
| IPF | J10_OTHERINSTPULM | 1 | 1 | 11_1219991_G_T | 11_1219991_G_T |
| IPF | R18_ABNORMAL_FINDI_DIAGNOST_IMAGI_LUNG | 1 | 1 | 11_1219991_G_T | 11_1219991_G_T |
| IPF | J10_INTERSTITIUM | 1 | 1 | 11_1219991_G_T | 11_1219991_G_T |
| IPF | ILD_INSUFFICIENCY | 1 | 1 | 11_1219991_G_T | 11_1219991_G_T |
| IPF | J10_OTHERINSTPULM | 1 | 1 | 5_1272247_G_A | 5_1272247_G_A |
| IPF | ILD_ENDPOINTS | 1 | 1 | 5_1272247_G_A | 5_1272247_G_A |
| IPF | ILD | 1 | 1 | 5_1272247_G_A | 5_1272247_G_A |
| IPF | J10_INTERSTITIUM | 1 | 1 | 5_1272247_G_A | 5_1272247_G_A |
| IPF | ILD_ENDPOINTS | 1 | 1 | 5_1279370_T_C | 5_1279370_T_C |
| IPF | ILD | 1 | 1 | 5_1279370_T_C | 5_1279370_T_C |
| IPF | J10_OTHERINSTPULM | 1 | 1 | 5_1279370_T_C | 5_1279370_T_C |
| IPF | J10_INTERSTITIUM | 0.99 | 1 | 5_1279370_T_C | 5_1279370_T_C |
| IPF | R18_ABNORMAL_FINDI_DIAGNOST_IMAGI_FUNCTION_STUDI_WO_DIAGNOSIS | 0.99 | 0.99 | 11_1219991_G_T | 11_1219991_G_T |
| IPF | CD2_BENIGN | 0.99 | 0.99 | 5_1272247_G_A | 5_1272247_G_A |
| IPF | ILD_INSUFFICIENCY | 0.98 | 0.98 | 5_1279370_T_C | 5_1279370_T_C |
| IPF | CD2_BENIGN_EXALLC | 0.98 | 0.98 | 5_1272247_G_A | 5_1272247_G_A |
| IPF | C3_SKIN | 0.83 | 0.9 | 5_169588475_G_A | 5_169588475_G_A |
| IPF | J10_INTERSTITIUM | 0.73 | 0.9 | 5_169588475_G_A | 5_169588475_G_A |
| IPF | C3_PROSTATE_EXALLC | 0.87 | 0.87 | 5_169588475_G_A | 5_169588475_G_A |
| IPF | C3_CANCER | 0.87 | 0.87 | 5_169588475_G_A | 5_169588475_G_A |
| IPF | C3_MALE_GENITAL_EXALLC | 0.87 | 0.87 | 5_169588475_G_A | 5_169588475_G_A |
| IPF | C3_PROSTATE | 0.87 | 0.87 | 5_169588475_G_A | 5_169588475_G_A |

| | | | | | |
|---|---|---|---|---|---|
| IPF | C3_MALE_GENITAL | 0.87 | 0.87 | 5_169588475_G_A | 5_169588475_G_A |
| IPF | C3_OTHER_SKIN_EXALLC | 0.86 | 0.87 | 5_169588475_G_A | 5_169588475_G_A |
| IPF | C3_BASAL_CELL_CARCINOMA_INCLAVO | 0.86 | 0.87 | 5_169588475_G_A | 5_169588475_G_A |
| IPF | C3_SKIN_EXALLC | 0.86 | 0.87 | 5_169588475_G_A | 5_169588475_G_A |
| IPF | C3_BASAL_CELL_CARCINOMA | 0.84 | 0.87 | 5_169588475_G_A | 5_169588475_G_A |
| IPF | C3_OTHER_SKIN | 0.84 | 0.87 | 5_169588475_G_A | 5_169588475_G_A |
| IPF | J10_INTERSTITIUM | 0.44 | 0.76 | 16_276685_G_A | 16_276685_G_A |
| IPF | OSTEOPOROSIS_FRACTURE_FG | 0.28 | 0.74 | 16_276685_G_A | 16_276685_G_A |
| IPF | CD2_LYMPHOID_LEUKAEMIA_EXALLC | 0.03 | 0.73 | 3_169759718_A_G | 3_169759718_A_G |
| IPF | CD2_LYMPHOID_LEUKAEMIA | 0.03 | 0.73 | 3_169759718_A_G | 3_169759718_A_G |
| IPF | J10_OTHERINSTPULM | 0.53 | 0.71 | 16_276685_G_A | 16_276685_G_A |
| IPF | ILD | 0.53 | 0.71 | 16_276685_G_A | 16_276685_G_A |
| IPF | ILD_ENDPOINTS | 0.53 | 0.71 | 16_276685_G_A | 16_276685_G_A |
| IPF | ILD_ENDPOINTS | 0.53 | 0.66 | 5_169588475_G_A | 5_169588475_G_A |
| IPF | ILD | 0.53 | 0.66 | 5_169588475_G_A | 5_169588475_G_A |
| IPF | ILD | 0.03 | 0.66 | 3_169759718_A_G | 3_169763483_C_G |
| IPF | ILD_ENDPOINTS | 0.03 | 0.66 | 3_169759718_A_G | 3_169763483_C_G |
| IPF | J10_OTHERINSTPULM | 0.53 | 0.66 | 5_169588475_G_A | 5_169588475_G_A |
| IPF | E4_HYTHY_AI_STRICT_PURCH | 0.0003 | 0.66 | 17_45753354_AGGAGTGAGCCGTGTGCGCATGGATGGGGGAAGGAGTGAGCCGTGTGCGCATGGATGGGGGAAGGAGTGAGCCGTGTGCGCATGGATGGGGGAGGGAGTGAGCCGTGTGCGCATGGATGGGGGAG_A | 17_46788132_C_T |
| IPF | M13_OSTEOPOROSIS | 0.2 | 0.58 | 16_276685_G_A | 16_276685_G_A |
| IPF | CD2_BENIGN_LEIOMYOMA_UTERI_EXALLC | 0.03 | 0.54 | 3_169759718_A_G | 3_169759718_A_G |
| IPF | CD2_BENIGN_LEIOMYOMA_UTERI | 0.03 | 0.49 | 3_169759718_A_G | 3_169759718_A_G |
| IPF | CD2_BENIGN_MELANOCYTIC | 0.02 | 0.48 | 3_169759718_A_G | 3_169774313_C_T |

| | | | | | |
|---|---|---|---|---|---|
| IPF | CD2_BENIGN_MELANOCYTIC_EXALLC | 0.02 | 0.47 | 3_169759718_A_G | 3_169769649_G_A |
| IPF | CD2_BENIGN_LIPOMATOUS_EXALLC | 0.02 | 0.46 | 3_169759718_A_G | 3_169774313_C_T |
| IPF | CD2_BENIGN_EXALLC | 0.02 | 0.27 | 3_169759718_A_G | 3_169769649_G_A |
| IPF | CD2_BENIGN | 0.02 | 0.25 | 3_169759718_A_G | 3_169769649_G_A |
| IPF | E4_HYTHYNAS | 0.0001 | 0.15 | 17_45753354_AGGAGTGAGCCGTGTGCGCATGGATGGGGGAAGGAGTGAGCCGTGTGCGCATGGATGGGGGAAGGAGTGAGCCGTGTGCGCATGGATGGGGGAGGGAGTGAGCCGTGTGCGCATGGATGGGGGAG_A | 17_46788132_C_T |
| IPF | HYPOT_THYRO_INCL | 0.0002 | 0.15 | 17_45753354_AGGAGTGAGCCGTGTGCGCATGGATGGGGGAAGGAGTGAGCCGTGTGCGCATGGATGGGGGAAGGAGTGAGCCGTGTGCGCATGGATGGGGGAGGGAGTGAGCCGTGTGCGCATGGATGGGGGAG_A | 17_46788132_C_T |
| IPF | C3_BREAST_EXALLC | 0.1 | 0.14 | 5_169588475_G_A | 5_169462850_C_G |
| IPF | C3_BREAST_HER2NEG_EXALLC | 0.06 | 0.1 | 5_169588475_G_A | 5_169462850_C_G |
| IPF | E4_DM2NOCOMP | 0.0018 | 0.03 | 11_1468491_G_A | 11_627276_C_T |

#### GBMI Biobank Acknowledgments

Biobank Japan Project

The BioBank Japan Project was supported by the Tailor-Made Medical Treatment program of the Ministry of Education, Culture, Sports, Science, and Technology (MEXT), the Japan Agency for Medical Research and Development (AMED). S.N. was supported by Takeda Science Foundation. Y.O. was supported by JSPS KAKENHI (19H01021, 20K21834), and AMED (JP21km0405211, JP21ek0109413, JP21ek0410075, JP21gm4010006, and JP21km0405217), JST Moonshot R&D (JPMJMS2021, JPMJMS2024), Takeda Science Foundation, and Bioinformatics Initiative of Osaka University Graduate School of Medicine, Osaka University.

BioMe - The Mount Sinai BioMe Biobank

The Mount Sinai BioMe Biobank has been supported by The Andrea and Charles Bronfman Philanthropies and in part by Federal funds from the NHLBI and NHGRI (U01HG00638001; U01HG007417; X01HL134588). We thank all participants in the Mount Sinai Biobank. We also thank all our recruiters who have assisted and continue to assist in data collection and management and are grateful for the computational resources and staff expertise provided by Scientific Computing at the Icahn School of Medicine at Mount Sinai.

BioVU

The BioVU projects at Vanderbilt University Medical Center are supported by numerous sources: institutional funding, private agencies, and federal grants. These include the NIH-funded Shared Instrumentation Grant S10OD017985 and S10RR025141; CTSA grants UL1TR002243, UL1TR000445, and UL1RR024975 from the National Center for Advancing Translational Sciences. Its contents are solely the responsibility of the authors and do not necessarily represent official views of the National Center for Advancing Translational Sciences or the National Institutes of Health. Genomic data are also supported by investigator-led projects that include U01HG004798, R01NS032830, RC2GM092618, P50GM115305, U01HG006378, U19HL065962, R01HD074711; and additional funding sources listed at https://victr. vumc.org/biovu-funding/.

Colorado Center for Personalized Medicine (CCPM)

The Colorado Center for Personalized Medicine (CCPM) would like to thank Richard Zane, Steve Hess, Sarah White, Emily Hearst, Emily Roberts and the entire Health Data Compass team. CCPM was developed with support from UCHealth, Children's Hospital Colorado, CU Medicine, CU Department of Medicine and CU School of Medicine.

China Kadoorie Biobank collaborative group

International Steering Committee: Junshi Chen, Zhengming Chen (PI), Robert Clarke, Rory Collins, Yu Guo, Liming Li (PI), Jun Lv, Richard Peto, Robin Walters, Chen Wang.

International Co-ordinating Centre, Oxford: Daniel Avery, Fiona Bragg, Derrick Bennett, Ruth Boxall, Ka Hung Chan, Yumei Chang, Yiping Chen, Zhengming Chen, Johnathan Clarke; Robert Clarke, Huaidong Du, Zammy Fairhurst-Hunter, Hannah Fry, Simon Gilbert, Alex Hacker, Parisa Hariri, Mike Hill, Michael Holmes, Pek Kei Im, Andri Iona, Maria Kakkoura, Christiana Kartsonaki, Rene Kerosi, Kuang Lin, Mohsen Mazidi, Iona Millwood, Qunhua Nie, Alfred Pozarickij, Paul Ryder, Sam Sansome, Dan Schmidt, Paul Sherliker, Rajani Sohoni, Becky Stevens, Iain Turnbull, Robin Walters, Lin Wang, Neil Wright, Ling Yang, Xiaoming Yang, Pang Yao. National Co-ordinating Centre, Beijing: Yu Guo, Xiao Han, Can Hou, Chun Li, Chao Liu, Jun Lv, Pei Pei, Canqing Yu.

Regional Coordinating Centres:

Guangxi Provincial CDC: Naying Chen, Duo Liu, Zhenzhu Tang. Liuzhou CDC: Ningyu Chen, Qilian Jiang, Jian Lan, Mingqiang Li, Yun Liu, Fanwen Meng, Jinhuai Meng, Rong Pan, Yulu Qin, Ping Wang, Sisi Wang, Liuping Wei, Liyuan Zhou. Gansu Provincial CDC: Caixia Dong, Pengfei Ge, Xiaolan Ren. Maiji CDC: Zhongxiao Li, Enke Mao, Tao Wang, Hui Zhang, Xi Zhang. Hainan Provincial CDC: Jinyan Chen, Ximin Hu, Xiaohuan Wang. Meilan CDC: Zhendong Guo, Huimei Li, Yilei Li, Min Weng, Shukuan Wu. Heilongjiang Provincial CDC: Shichun Yan, Mingyuan Zou, Xue Zhou. Nangang CDC: Ziyan Guo, Quan Kang, Yanjie Li, Bo Yu, Qinai Xu. Henan Provincial CDC: Liang Chang, Lei Fan, Shixian Feng, Ding Zhang, Gang Zhou. Huixian CDC: Yulian Gao, Tianyou He, Pan He, Chen Hu, Huarong Sun, Xukui Zhang. Hunan Provincial CDC: Biyun Chen, Zhongxi Fu, Yuelong Huang, Huilin Liu, Qiaohua Xu, Li Yin. Liuyang CDC: Huajun Long, Xin Xu, Hao Zhang, Libo Zhang. Jiangsu Provincial CDC: Jian Su, Ran Tao, Ming Wu, Jie Yang, Jinyi Zhou, Yonglin Zhou. Suzhou CDC: Yihe Hu, Yujie Hua, Jianrong Jin Fang Liu, Jingchao Liu, Yan Lu, Liangcai Ma, Aiyu Tang, Jun Zhang. Qingdao CDC: Liang Cheng, Ranran Du, Ruqin Gao, Feifei Li, Shanpeng Li, Yongmei Liu, Feng Ning, Zengchang Pang, Xiaohui Sun, Xiaocao Tian, Shaojie Wang, Yaoming Zhai, Hua Zhang. Licang CDC: Wei Hou, Silu Lv, Junzheng Wang. Sichuan Provincial CDC: Xiaofang Chen, Xianping Wu, Ningmei Zhang, Weiwei Zhou. Pengzhou CDC: Xiaofang Chen, Jianguo Li, Jiaqiu Liu, Guojin Luo, Qiang Sun, Xunfu Zhong. Zhejiang Provincial CDC: Weiwei Gong, Ruying Hu, Hao Wang,Meng Wan, Min Yu. Tongxiang CDC: Lingli Chen, Qijun Gu, Dongxia Pan, Chunmei Wang, Kaixu Xie, Xiaoyi Zhang.

CKB Acknowledgements and Funding:

China Kadoorie Biobank gratefully acknowledges the participants, project staff, and the China National Centre for Disease Control and Prevention (CDC) and its regional offices. China's National Health Insurance provides electronic linkage to all hospital treatment. Funding sources: Baseline survey and first re-survey – Kadoorie Charitable Foundation, Hong Kong; long-term follow-up – UK Wellcome Trust (212946/Z/18/Z, 202922/Z/16/Z, 104085/Z/14/Z, 088158/Z/09/Z), National Natural Science Foundation of China (91843302), National Key Research and Development Program of China (2016YFC 0900500, 0900501, 0900504, 1303904); DNA extraction and genotyping – GlaxoSmithKline, UK Medical Research Council (MC-PC-13049, MC-PC-14135); core funding for the project to the Clinical Trial Service Unit and Epidemiological Studies Unit at Oxford University – British Heart Foundation (CH/1996001/9454), UK Medical Research Council (MC-UU-00017/1, MC-UU-12026/2, MC_U137686851), Cancer Research UK (C16077/A29186, C500/A16896).

Estonian Biobank
This research was supported by the European Union through Horizon 2020 research and innovation programme under grant no 810645 and through the European Regional Development Fund project no. MOBEC008, by the Estonian Research Council grant PUT (PRG1291, PRG687 and PRG184) and by the European Union through the European Regional Development Fund project no. MOBERA21 (ERA-CVD project DETECT ARRHYTHMIAS, GA no JTC2018-009), Project No. 2014-2020.4.01.15-0012 and Project No. 2014-2020.4.01.16-0125. We would like to acknowledge Dr. Tõnu Esko; Dr. Lili Milani; Dr. Reedik Mägi, Dr. Mari Nelis and Dr. Andres Metspalu, all from the Institute of Genomics, University of Tartu, Tartu, Estonia.

FinnGen
The FinnGen project is funded by two grants from Business Finland (HUS 4685/31/2016 and UH 4386/31/2016) and the following industry partners: AbbVie Inc., AstraZeneca UK Ltd, Biogen MA Inc., Bristol Myers Squibb (and Celgene Corporation & Celgene International II Sàrl), Genentech Inc., Merck Sharp & Dohme Corp, Pfizer Inc., GlaxoSmithKline Intellectual Property Development Ltd., Sanofi US Services Inc., Maze Therapeutics Inc., Janssen Biotech Inc, and Novartis AG. Following biobanks are acknowledged for delivering biobank samples to FinnGen: Auria Biobank (www.auria.fi/biopankki), THL Biobank (www.thl.fi/biobank), Helsinki Biobank (www.helsinginbiopankki.fi), Biobank Borealis of Northern Finland (https://www.ppshp.fi/Tutkimus-ja-opetus/Biopankki/Pages/Biobank-Borealis-briefly-in-English.aspx), Finnish Clinical Biobank Tampere (www.tays.fi/en-US/Research_and_development/Finnish_Clinical_Biobank_Tampere), Biobank of Eastern Finland (www.ita-suomenbiopankki.fi/en), Central Finland Biobank (www.ksshp.fi/fi-FI/Potilaalle/Biopankki), Finnish Red Cross Blood Service Biobank (www.veripalvelu.fi/verenluovutus/biopankkitoiminta) and Terveystalo Biobank (www.terveystalo.com/fi/Yritystietoa/Terveystalo-Biopankki/Biopankki/). All Finnish Biobanks are members of BBMRI.fi infrastructure (www.bbmri.fi). Finnish Biobank Cooperative -FINBB (https://finbb.fi/) is the coordinator of BBMRI-ERIC operations in Finland. The Finnish biobank data can be accessed through the Fingenious® services (https://site.fingenious.fi/en/) managed by FINBB.

Genes and Health
Genes & Health is/has recently been core-funded by Wellcome (WT102627, WT210561), the Medical Research Council (UK) (M009017), Higher Education Funding Council for England Catalyst, Barts Charity (845/1796), Health Data Research UK (for London substantive site), and research delivery support from the NHS National Institute for Health Research Clinical

Research Network (North Thames). We thank Social Action for Health, Centre of The Cell, members of our Community Advisory Group, and staff who have recruited and collected data from volunteers. We thank the NIHR National Biosample Centre (UK Biocentre), the Social Genetic & Developmental Psychiatry Centre (King's College London), Wellcome Sanger Institute, and Broad Institute for sample processing, genotyping, sequencing and variant annotation. We thank: Barts Health NHS Trust, NHS Clinical Commissioning Groups (City and Hackney, Waltham Forest, Tower Hamlets, Newham, Redbridge, Havering, Barking and Dagenham), East London NHS Foundation Trust, Bradford Teaching Hospitals NHS Foundation Trust, Public Health England (especially David Wyllie), Discovery Data Service/Endeavour Health Charitable Trust (especially David Stables) - for GDPR-compliant data sharing backed by individual written informed consent. Most of all we thank all of the volunteers participating in Genes & Health.
Genes & Health Research Team (in alphabetical order by surname): Shaheen Akhtar, Mohammad Anwar, Elena Arciero, Samina Ashraf, Gerome Breen, Raymond Chung, Charles J Curtis, Maharun Chowdhury, Grainne Colligan, Panos Deloukas, Ceri Durham, Sarah Finer, Chris Griffiths, Qin Qin Huang, Matt Hurles, Karen A Hunt, Shapna Hussain, Kamrul Islam, Ahsan Khan, Amara Khan, Cath Lavery, Sang Hyuck Lee, Robin Lerner, Daniel MacArthur, Bev MacLaughlin, Hilary Martin, Dan Mason, Shefa Miah, Bill Newman, Nishat Safa, Farah Tahmasebi, Richard C Trembath, Bhavi Trivedi, David A van Heel, John Wright.

The HUNT Study
A special thanks to all the HUNT participants for donating their time, samples and information to help others.
The Trøndelag Health Study (HUNT) is a collaboration between HUNT Research Centre (Faculty of Medicine and Health Sciences, NTNU, Norwegian University of Science and Technology), Trøndelag County Council, Central Norway Regional Health Authority, and the Norwegian Institute of Public Health. The genotyping in HUNT was financed by the National Institutes of Health; University of Michigan; the Research Council of Norway; the Liaison Committee for Education, Research and Innovation in Central Norway; and the Joint Research Committee between St Olavs hospital and the Faculty of Medicine and Health Sciences, NTNU. The genetic investigations of the HUNT Study, is a collaboration between researchers from the K.G. Jebsen Center for Genetic Epidemiology, NTNU and the University of Michigan Medical School and the University of Michigan School of Public Health. The K.G. Jebsen Center for Genetic Epidemiology is financed by Stiftelsen Kristian Gerhard Jebsen; Faculty of Medicine and Health Sciences, NTNU, Norway.
We want to thank clinicians and other employees at Nord-Trøndelag Hospital Trust for their support and for contributing to data collection in this research project.
We also acknowledge; HUNT-MI Leadership: Kristian Hveem, Cristen Willer, Oddgeir Lingaas Holmen, Mike Boehnke, Goncalo Abecasis, Bjorn Olav Åsvold, Ben Brumpton; Scientific Advisory Committee: Ele Zeggini, Mark Daly, Bjørn Pasternak; HUNT Research Centre: Jørn

Søberg Fenstad, Anne Jorunn Vikdal, Marit Næss; HUNT Cloud: Oddgeir Lingaas Holmen, Sandor Zeestraten, Tom Erik Røberg; Data applications and registry linkages: Maiken E. Gabrielsen, Anne Heidi Skogholt; Low-pass whole sequencing genome bioinformatics and statistical analysis: He Zhang, Hyun Min Kang, Jin Chen; Array genotyping: Sten Even Erlandsen, Vidar Beisvåg; GWAS bioinformatics, QC, imputation and statistical analysis: Wei Zhou, Jonas Nielsen, Lars Fritsche, Hyun Min Kang, Oddgeir Holmen, Ben Brumpton, Laurent Thomas; CNV calling: Ellen Schmidt, Ryan Mills; Statistical methods development for analyzing HUNT data: Wei Zhou, Shawn Lee
Funding:
The K. G. Jebsen Centre for Genetic Epidemiology is financed by Stiftelsen Kristian Gerhard Jebsen. The genotyping in HUNT was financed by the National Institutes of Health; University of Michigan; the Research Council of Norway; Stiftelsen Kristian Gerhard Jebsen; the Liaison Committee for Education, Research and Innovation in Central Norway; and the Joint Research Committee between St Olav's hospital and the Faculty of Medicine and Health Sciences, NTNU.

Mass General Brigham (MGB) Biobank
Samples, genomic data, and health information were obtained from the Mass General Brigham Biobank, a biorepository of consented patients samples at Mass General Brigham (parent organization of Massachusetts General Hospital and Brigham and Women's Hospital). We are grateful to all of the participants and clinical and research teams who made this work possible. Support for genotyping was provided through MGB Personalized Medicine.
MGB Biobank Leadership: Elizabeth W. Karlson, MD; Shawn N. Murphy, MD, PhD; Susan A Slaugenhaupt, PhD; Jordan W. Smoller, MD, ScD; Scott T. Weiss, MD, MSc

Michigan Genomics Initiative
The authors acknowledge the Michigan Genomics Initiative participants, Precision Health at the University of Michigan, the University of Michigan Medical School Central Biorepository, and the University of Michigan Advanced Genomics Core for providing data and specimen storage, management, processing, and distribution services, and the Center for Statistical Genetics in the Department of Biostatistics at the School of Public Health for genotype data curation, imputation, and management in support of the research reported in this publication.

UCLA ATLAS Community Health Initiative (UCLA)
We gratefully acknowledge the resources provided by the Institute for Precision Health (IPH) and participating UCLA ATLAS Community Health Initiative patients. The UCLA ATLAS Community Health Initiative in collaboration with UCLA ATLAS Precision Health Biobank, is a program of IPH, which directs and supports the biobanking and genotyping of biospecimen samples from participating UCLA patients in collaboration with the David Geffen School of Medicine, UCLA CTSI and UCLA Health. Members of the UCLA ATLAS Community Health

Initiative include Ruth Johnson, Yi Ding, Vidhya Venkateswaran, Arjun Bhattacharya, Alec Chiu, Tommer Schwarz, Malika Freund, Lingyu Zhan, Kathryn S. Burch, Christa Caggiano, Brian Hill, Nadav Rakocz, Brunilda Balliu, Jae Hoon Sul, Noah Zaitlen, Valerie A. Arboleda, Eran Halperin, Sriram Sankararaman, Manish J. Butte, Clara Lajonchere, Daniel H. Geschwind, and Bogdan Pasaniuc, on behalf of theUCLA Precision Health Data Discovery Repository Working Group and UCLA Precision Health ATLAS Working Group.

UK Biobank
Access to data from the UK BioBank was obtained through Application #31063
PI: Ben Neale, Claire Churchhouse
Overview: “Methodological extensions to estimate genetic heritability and shared risk factors for phenotypes of the UK Biobank”.
Website for Pan-UKBB results can be found: https://pan.ukbb.broadinstitute.org/

Other
G.D.S.; T.R.G.; and J.Z. are supported by a grant from the Medical Research Council for the Integrative Epidemiology Unit at the University of Bristol MC_UU_00011/1 & 4.
J.Z. is supported by the Academy of Medical Sciences (AMS) Springboard Award, the Wellcome Trust, the Government Department of Business, Energy and Industrial Strategy (BEIS), the British Heart Foundation and Diabetes UK (SBF006\1117). J.Z. is funded by the Vice-Chancellor Fellowship from the University of Bristol. W.Z. was supported by the National Human Genome Research Institute of the National Institutes of Health under award number T32HG010464.

ICDA
The authors would like to acknowledge the organizing committee of the International Common Disease Alliance for intellectual contributions on the set up of the GBMI as a nascent activity to the larger effort. We also thank them for the use of their slack platform.
Website for ICDA can be found here: https://www.icda.bio/

The Hail Team and Data Management at the Stanley Center for Psychiatric Research
Hail is an open-source Python library that simplifies genomic data analysis in the cloud. It provides powerful, easy-to-use data science tools that can be used to interrogate biobank-scale genomic data and was used in the analysis of the data for this paper. We would especially like to thank Daniel King from the Hail team and Sam Bryant from the Stanley Center Data Management team for helping with the Google bucket set up and data sharing.
Website for Hail can be found here: https://hail.is/

#### GBMI authors

Wei Zhou1,2,3, Masahiro Kanai1,2,3,4,5, Kuan-Han H Wu6, Humaira Rasheed7,8,9, Kristin Tsuo1,2,3, Jibril B Hirbo10,11, Ying Wang1,2,3, Arjun Bhattacharya12, Huiling Zhao9, Shinichi Namba5, Ida Surakka13, Brooke N Wolford6,7, Valeria Lo Faro14,15,16, Esteban A Lopera-Maya17, Kristi Läll18, Marie-Julie Favé19, Sinéad B Chapman2,3, Juha Karjalainen1,2,3,20, Mitja Kurki1,2,3,20, Maasha Mutaamba1,2,3,20, Juulia Partanen20, Ben M Brumpton7,21,22, Sameer Chavan23, Tzu-Ting Chen24, Michelle Daya23, Yi Ding12,25, Yen-Chen A Feng26,27, Christopher R Gignoux23, Sarah E Graham13, Whitney E Hornsby13, Nathan Ingold28,29, Ruth Johnson12,30, Triin Laisk18, Kuang Lin31, Jun Lv32, Iona Y Millwood31,33, Priit Palta18,20, Anita Pandit34, Michael H Preuss35, Unnur Thorsteinsdottir36, Jasmina Uzunovic19, Matthew Zawistowski34, Xue Zhong10,11, Archie Campbell37, Kristy Crooks23, Geertruida H de Bock38, Nicholas J Douville39,40, Sarah Finer41, Lars G Fritsche34, Christopher J Griffiths41, Yu Guo42, Karen A Hunt43, Takahiro Konuma5,44, Riccardo E Marioni37, Jansonius Nomdo14, Snehal Patil34, Nicholas Rafaels23, Anne Richmond45, Jonathan A Shortt23, Peter Straub10,11, Ran Tao11, Brett Vanderwerff34, Kathleen C Barnes23, Marike Boezen‡38, Zhengming Chen31,33, Chia-Yen Chen46, Judy Cho35, George Davey Smith9,47, Hilary K Finucane1,2,3, Lude Franke17, Eric R Gamazon10,11,48, Andrea Ganna1,2,20, Tom R Gaunt9, Tian Ge27,49, Hailiang Huang1,2, Jennifer Huffman50, Jukka T. Koskela20, Clara Lajonchere51,52, Matthew H Law28,29, Liming Li32, Cecilia M Lindgren53, Ruth JF Loos35,54, Stuart MacGregor28, Koichi Matsuda55, Catherine M Olsen28, David J Porteous37, Jordan A Shavit56, Harold Snieder38, Richard C Trembath57, Judith M Vonk38, David Whiteman28, Stephen J Wicks23, Cisca Wijmenga17, John Wright58, Jie Zheng9, Xiang Zhou34, Philip Awadalla19,59, Michael Boehnke34, Nancy J Cox10,11, Daniel H Geschwind51,60,61, Caroline Hayward45, Kristian Hveem7,21, Eimear E Kenny62, Yen-Feng Lin24,63,64, Reedik Mägi18, Hilary C Martin65, Sarah E Medland28, Yukinori Okada5,66,67,68,69, Aarno V Palotie1,2,20, Bogdan Pasaniuc12,25,51,60,70, Serena Sanna17,71, Jordan W Smoller27, Kari Stefansson36, David A van Heel43, Robin G Walters31,33, Sebastian Zöllner34, Biobank Japan, BioMe, BioVU, Canadian Partnership for Tomorrow's Health/Ontario Health Study, China Kadoorie Biobank Collaborative Group, Colorado Center for Personalized Medicine, deCODE Genetics, Estonian Biobank, FinnGen, Generation Scotland, Genes & Health, LifeLines, Mass General Brigham Biobank, Michigan Genomics Initiative, QIMR Berghofer Biobank, Taiwan Biobank, The HUNT Study, UCLA ATLAS Community Health Initiative, UK Biobank, Alicia R Martin1,2,3, Cristen J Willer6,13,72*, Mark J Daly1,2,3,20*, Benjamin M Neale1,2,3*

‡Deceased

*These authors jointly supervised this work

1Analytic and Translational Genetics Unit, Department of Medicine, Massachusetts General Hospital, Boston, MA, USA, 2Stanley Center for Psychiatric Research, Broad Institute of MIT and Harvard, Cambridge, MA, USA, 3Program in Medical and Population Genetics, Broad

Institute of MIT and Harvard, Cambridge, MA, USA, 4Department of Biomedical Informatics, Harvard Medical School, Boston, MA, USA, 5Department of Statistical Genetics, Osaka University Graduate School of Medicine, Suita 565-0871, Japan, 6Department of Computational Medicine and Bioinformatics, University of Michigan, Ann Arbor, MI, USA, 7K.G. Jebsen Center for Genetic Epidemiology, Department of Public Health and Nursing, NTNU, Norwegian University of Science and Technology, Trondheim, Norway, 8Division of Medicine and Laboratory Sciences, University of Oslo, Norway, 9MRC Integrative Epidemiology Unit (IEU), Bristol Medical School, University of Bristol, Bristol, UK, 10Department of Medicine, Division of Genetic Medicine, Vanderbilt University Medical Center, Nashville, TN, USA, 11Vanderbilt Genetics Institute, Vanderbilt University Medical Center, Nashville, TN, USA, 12Department of Pathology and Laboratory Medicine, David Geffen School of Medicine, University of California, Los Angeles, Los Angeles, CA, USA, 13Department of Internal Medicine, Division of Cardiology, University of Michigan, Ann Arbor, MI, USA, 14University of Groningen, UMCG, Department of Ophthalmology, Groningen, the Netherlands, 15Department of Clinical Genetics, Amsterdam University Medical Center (AMC), Amsterdam, the Netherlands, 16Department of Immunology, Genetics and Pathology, Science for Life Laboratory, Uppsala University, Uppsala, Sweden, 17University of Groningen, UMCG, Department of Genetics, Groningen, the Netherlands, 18Estonian Genome Centre, Institute of Genomics, University of Tartu, Tartu, Estonia, 19Ontario Institute for Cancer Research, Toronto, ON, Canada, 20Institute for Molecular Medicine Finland, University of Helsinki, Helsinki, Finland, 21HUNT Research Centre, Department of Public Health and Nursing, NTNU, Norwegian University of Science and Technology, Levanger, Norway, 22Clinic of Medicine, St. Olavs Hospital, Trondheim University Hospital, Trondheim, Norway, 23University of Colorado - Anschutz Medical Campus, Aurora, CO, USA, 24Center for Neuropsychiatric Research, National Health Research Institutes, Miaoli, Taiwan, 25Bioinformatics Interdepartmental Program, University of California, Los Angeles, Los Angeles, CA, USA, 26Division of Biostatistics, Institute of Epidemiology and Preventive Medicine, College of Public Health, National Taiwan University, Taiwan, 27Psychiatric and Neurodevelopmental Genetics Unit, Center for Genomic Medicine, Massachusetts General Hospital, Boston, MA, USA, 28QIMR Berghofer Medical Research Institute, Brisbane, Australia, 29Faculty of Health, School of Biomedical Sciences, Queensland University of Technology, Australia, 30Department of Computer Science, University of California, Los Angeles, Los Angeles, CA, USA, 31Nuffield Department of Population Health, University of Oxford, Oxford, UK, 32Department of Epidemiology and Biostatistics, School of Public Health, Peking University Health Science Center, Beijing, China, 33MRC Population Health Research Unit, University of Oxford, Oxford, UK, 34Department of Biostatistics and Center for Statistical Genetics, University of Michigan, Ann Arbor, MI, USA, 35The Charles Bronfman Institute for Personalized Medicine, Icahn School of Medicine at Mount Sinai, New York, NY, USA, 36deCODE Genetics/Amgen inc., 101, Reykjavik, Iceland, 37Centre for Genomic and Experimental Medicine, Institute of Genetics and Cancer, University of Edinburgh, Edinburgh, UK, 38University of Groningen, UMCG, Department of Epidemiology, Groningen, the

Netherlands, 39Department of Anesthesiology, Michigan Medicine, Ann Arbor, MI, USA, 40Institute of Healthcare Policy & Innovation, University of Michigan, Ann Arbor, MI, USA, 41Wolfson Institute of Population Health, Queen Mary University of London, London, UK, 42Chinese Academy of Medical Sciences, Beijing, China, 43Blizard Institute, Queen Mary University of London, London, UK, 44Central Pharmaceutical Research Institute, JAPAN TOBACCO INC., Takatsuki 569-1125, Japan, 45Medical Research Council Human Genetics Unit, Institute of Genetics and Cancer, University of Edinburgh, Edinburgh, UK, 46Biogen, Cambridge, MA, USA, 47NIHR Bristol Biomedical Research Centre, Bristol, UK, 48MRC Epidemiology Unit, University of Cambridge, Cambridge, UK, 49Center for Precision Psychiatry, Massachusetts General Hospital, Boston, MA, USA, 50Centre for Population Genomics, VA Boston Healthcare System, Boston, MA, USA, 51Institute of Precision Health, University of California, Los Angeles, Los Angeles, CA, USA, 52Program in Neurogenetics, Department of Neurology, David Geffen School of Medicine, University of California, Los Angeles, Los Angeles, CA, USA, 53Big Data Institute, Li Ka Shing Centre for Health Information and Discovery, University of Oxford, Oxford, UK, 54Novo Nordisk Foundation Center for Basic Metabolic Research, Faculty of Medicine and Health Sciences, University of Copenhagen, Copenhagen, Denmark, 55Department of Computational Biology and Medical Sciences, Graduate school of Frontier Sciences, The University of Tokyo, Tokyo, Japan, 56University of Michigan, Department of Pediatrics, Ann Arbor MI 48109, 57School of Basic and Medical Biosciences, Faculty of Life Sciences and Medicine, King's College London, London, UK, 58Bradford Institute for Health Research, Bradford Teaching Hospitals National Health Service (NHS) Foundation Trust, Bradford, UK, 59Department of Molecular Genetics, University of Toronto, Toronto, ON, Canada, 60Department of Human Genetics, David Geffen School of Medicine, University of California, Los Angeles, Los Angeles, CA, USA, 61Department of Neurology, David Geffen School of Medicine, University of California, Los Angeles, Los Angeles, CA, USA, 62Institute for Genomic Health, Icahn School of Medicine at Mount Sinai, New York, NY, USA, 63Department of Public Health & Medical Humanities, School of Medicine, National Yang Ming Chiao Tung University, Taipei, Taiwan, 64Institute of Behavioral Medicine, College of Medicine, National Cheng Kung University, Tainan, Taiwan, 65Medical and Population Genomics, Wellcome Sanger Institute, Hinxton, UK, 66Center for Infectious Disease Education and Research (CiDER), Osaka University, Suita 565-0871, Japan, 67Laboratory of Statistical Immunology, Immunology Frontier Research Center (WPI-IFReC), Osaka University, Suita 565-0871, Japan, 68Laboratory for Systems Genetics, RIKEN Center for Integrative Medical Sciences, Yokohama, Japan, 69Integrated Frontier Research for Medical Science Division, Institute for Open and Transdisciplinary Research Initiatives, Osaka University, Suita 565-0871, Japan, 70Department of Computational Medicine, David Geffen School of Medicine, University of California, Los Angeles, Los Angeles, CA, USA, 71Institute for Genetics and Biomedical Research (IRGB), National Research Council (CNR), Cagliari, Italy, 72Department of Human Genetics, University of Michigan, Ann Arbor, MI, USA
